# Prognostic and Diagnostic Utility of Heart Rate Variability to Predict and Understand Change in Cancer and Chemotherapy Related Fatigue, Pain, and Neuropathic Symptoms: A Systematic Review

**DOI:** 10.1101/2025.01.08.25320191

**Authors:** Jessica Bolanos, Layal Hneiny, Juan Gonzalez, Max O’Malley, Marlon L Wong

## Abstract

Advances in early cancer detection and treatment have significantly improved survival rates, resulting in over 18.1 million cancer survivors in the United States. Many of these survivors experience chronic pain, fatigue, and neuropathic symptoms related to cancer or its treatments. Emerging evidence suggests that autonomic nervous system dysfunction plays a crucial role in these symptoms. Heart rate variability (HRV), a measure of autonomic function, has shown potential in predicting the onset of somatic symptoms in cancer patients. This systematic review aimed to assess the association of HRV with pain, fatigue, and neuropathy in cancer patients and survivors.

A comprehensive search was conducted across multiple databases, yielding 19 studies that met inclusion criteria. These studies varied in cancer types, stages, and HRV measurement methods. Most studies focused on breast cancer and reported a predominant female population. Fatigue was the most studied symptom, followed by pain. HRV measures included both time and frequency domain variables, with significant variability in measurement duration and control for confounding factors.

Findings suggest that decreased HRV is associated with increased fatigue and pain, providing potential support for a bidirectional relationship between autonomic dysfunction and these symptoms. However, the heterogeneity in HRV measurement methods and the high risk of bias in many studies highlight the need for standardized HRV protocols in cancer research. Further large-scale studies with low risk of bias are necessary to validate HRV as a reliable tool for phenotyping and managing cancer-related symptoms.

## Introduction

Advances in early detection and treatment has significantly improved 5-year cancer-specific survival rates (1). As a result, there is currently over 18.1 million cancer survivors in the United States (1), many of whom suffer from chronic pain (2), fatigue (3), or neuropathic symptoms (i.e., paresthesia and dysesthesia) associated with cancer or cancer treatments (4, 5). There is growing evidence that dysfunction of the autonomic nervous system plays a key role in the development and maintenance of these comorbid symptoms (6–8). The autonomic nervous system is an important regulator of stress responses, and it exhibits functional changes in chemotherapy induced peripheral neuropathy (CIPN) and in other chronic pain conditions (8, 9). The vagus nerve is the primary nerve of the parasympathetic system, and research indicates that the vagus nerve modulates pain through 3 pathways: 1) by maintaining autonomic and hypothalamic-pituitary-adrenal axis balance thereby reducing allostatic load (10), 2) via the cholinergic anti-inflammatory pathway in which action potentials in the vagus nerve inhibit production of TNF and other inflammatory cytokines (11), and 3) by modulating the function of brain networks involving the cingulate cortex and insula which are involved in pain processing (12). Dysfunction of these pathways may also contribute to cancer related fatigue and neuropathy (8, 9, 13).

Measures of heart rate variability (HRV) are commonly used as surrogate measures of autonomic function, and emerging evidence suggests that changes in HRV may predict the onset of somatic symptoms in cancer patients. In fact, a recent study found that changes in HRV preceded the onset of CIPN symptoms in people receiving oxaliplatin for gastrointestinal cancer (8), and another study found that changes in HRV were correlated to sensory disturbance in people with CIPN (9). Decreased HRV has also been observed in other chronic pain populations, and it has been proposed that there is a bidirectional relationship between chronic pain and autonomic dysfunction (14). Therefore, HRV is a promising measurement tool for improving the phenotyping and measurement of cancer related somatic symptoms, and it is potentially a useful treatment target.

The purpose of this study was to assess the association of HRV with comorbid symptoms (i.e., pain, fatigue, and neuropathy) in cancer patients and survivors. However, the use and interpretation of HRV lacks standardization in research. Thus, this review also describes the methods used for HRV measurement in cancer research and discusses how these methods may influence findings.

## Materials and Methods

A systematic review was conducted following the Methodological Expectations of Cochrane Intervention Reviews Manual (15), and the results were then reported according to the Preferred Reporting Items for Systematic Reviews and Meta-analyses (PRISMA) (16). The review methods were established before developing the review, and the study protocol was registered at the International Prospective Register of Systematic Reviews under CRD42023418747.

### Search Strategy

The study inclusion criteria consisted of 1) studies including cancer patients or survivors, and 2) longitudinal cohort studies and randomized control trials (RCTs) that include at least 1 HRV measure timepoint and must include an objective measure of change for at least 1 of the symptoms listed (pain, fatigue, and/or neuropathy) or 3) cross-sectional studies that compare HRV between groups of cancer survivors with and without one of the symptoms of interest and allow for calculation of effect sizes. A search strategy was built by the clinical research librarian (LH) from Louis Calder Memorial Library together with the Primary Investigator (MW). The medical librarian (DG) reviewed it and gave an input using the Peer Review for Electronic Search Strategies tool (17).

Firstly, the search strategy was developed in the database Ovid using both the Medical Subject Headings for the concepts of Heart Rate Variability, Pain and Fatigue, and cancer and chemotherapy and their alternate keywords. In addition to Ovid MEDLINE(R) ALL, the following databases were searched: Cochrane Central Register of Controlled Trials (CENTRAL) (Cochrane Library, Wiley), Embase (Elsevier, Embase.com), Cumulative Index to Nursing and Allied Health Literature (CINAHL) Plus (Ebsco), the Web of Science platform (Clarivate: Science Citation Index Expanded, Social Sciences Citation Index, Arts & Humanities Citation Index, Conference Proceedings Citation Index-Science, Conference Proceedings Citation Index-Social Science & Humanities, Emerging Sources Citation Index), Scopus (Elsevier), PsycINFO, ProQuest Databases (ABI/INFORM Collection, Academic Video Online, AFI Catalog, American Periodicals, ASFA: Aquatic Sciences and Fisheries Abstracts, Black Studies Center, Business Market Research Collection, Coronavirus Research Database, Digital National Security Archive, Dissertations & Theses @ University of Miami, Early Modern Books, Ebook Central, ERIC, Ethnic NewsWatch, GenderWatch, Global Newsstream, Healthcare Administration Database, Music Periodicals Database, ProQuest Civil War Era, ProQuest Dissertations & Theses Global, ProQuest Historical Newspapers: Chicago Defender, ProQuest Historical Newspapers: New York Amsterdam News, ProQuest Historical Newspapers: New York Tribune, ProQuest Historical Newspapers: Philadelphia Tribune, ProQuest Historical Newspapers: The Baltimore Afro-American, ProQuest Historical Newspapers: The New York Times with Index, PTSDpubs, Publicly Available Content Database, Research Library, SciTech Premium Collection, Sociological Abstracts, Teatro Español del Siglo de Oro, Worldwide Political Science Abstracts) limited to Dissertations & Theses, and APA PsycInfo (Ebsco).

The Medline search strategy was adapted for the other databases each according to its syntax taking into consideration the use of different controlled vocabulary such as: EMTREE terms for Embase, Medical Subject Headings for Cochrane Library, and CINAHL Subject Headings for CINAHL plus, in addition to revising the truncation, adjacency, and search fields. No application for language, date, or other limitations was applied. All databases were searched on 8,9, and 10 November 2023. The full search strategy is available in Appendix 1.

### Study selection, data extraction, and data management

The results that were yielded from the databases were exported into Covidence Web-based software (18) directly or into EndNote 20 (19) then imported to Covidence. In Covidence, the duplicates of overlapping results were removed, then the remaining results were first screened for title-abstract, then screened for full-text (18). Each step was conducted by independently by two reviewers (JB and JG), and conflicts were resolved by consensus with the primary investigator (MW).

An Excel spreadsheet was developed and piloted for data extraction. Then, two independent reviewers (JB and MO) extracted data from the full-text manuscripts. Missing data was requested from the study authors via email. Seven authors were contacted in November 2024 via email requesting clarification of measures or raw data sets for symptom of interest and heart rate variability measures. Two authors responded with clarifications, two contacts were listed as undeliverable, and three authors did not respond.

### Risk of bias assessment and synthesis of findings

Our intention was to include all longitudinal studies with repeated measures of HRV and symptoms, which would likely encompass a variety of study designs. Thus, the Mixed Methods Appraisal Tool (MMAT) was chosen for risk of bias assessment due to its applicability to a wide range of study types (20). Each study was assessed by two independent reviewers (JG and MO) for risk of bias, and disagreements between reviewers were resolved by deliberation until a consensus is reached (Appendix 2).

The body of evidence was assessed based on the Cochrane GRADE classification (high, moderate, low and very low) (21), and an Excel spreadsheet was used to record the data. Four investigators (JB, JG, MO, and MW) deliberated until consensus was achieved for Cochrane GRADE classification.

## Results

### Study Characteristics

A total of 6677 titles and abstracts were screened by two reviewers. Fifty-nine full text articles were assessed with 19 articles ultimately selected for inclusion (**Figure 1**). Publication dates ranged from 2009-2023, and the studies covered a wide range of cancer types (**Table 1**). Nine of the 19 studies focused solely on people with breast cancer (22–30). Of the 19 studies, nine (26, 28, 30–36) did not report staging information, however most that reported staging ranged from stage 0 to terminal with most participants in stage I.

**Figure 1.**
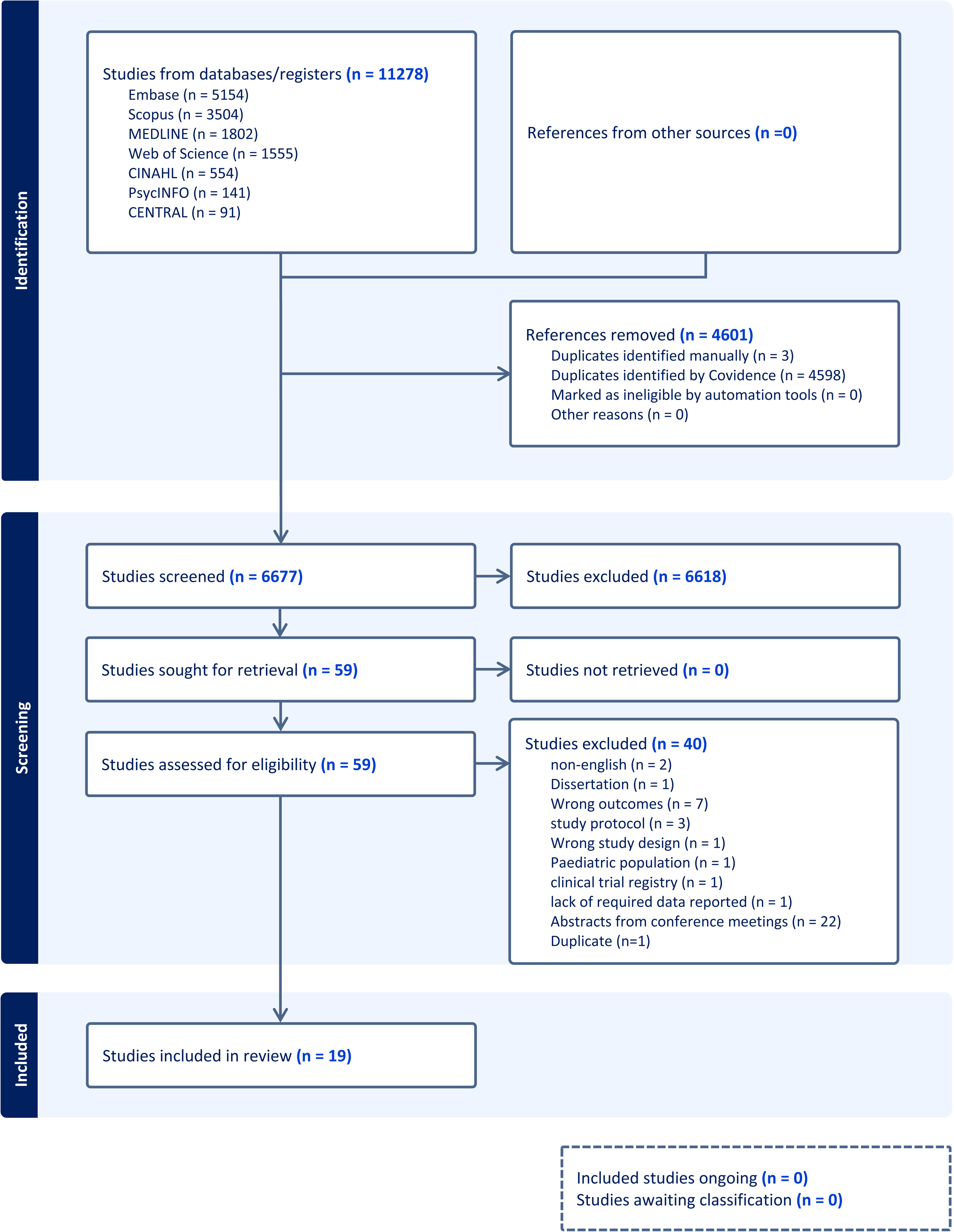
PRISMA Flow Diagram.

**Table 1.** Study Characteristics.

| First author (year) | Country | Aim of study | Design | Participants recruited | Heart rate variability measurement details | Key relevant findings |
| --- | --- | --- | --- | --- | --- | --- |
| Lee (2018) | Taiwan | To assess the effects of 12 weeks of Qigong exercise or stress management training on fatigue, fear of recurrence, quality of life, and heart rate variability in people who completed cancer treatment at least 1 month prior. | Randomized control trial | 99 cancer survivors | 5-minute recording seated or lying prone; PPG device; time domain as primary | After 12 weeks of both Qigong and stress management training, fatigue was reduced and increases in SDNN, TP and HF were observed. |
| Burch (2020) | USA | To assess if heart rate variability biofeedback treatment among cancer survivors can increase HRV coherence and reduce symptoms of pain, stress, fatigue, depression, distress, PTSD, and insomnia relative to control patients in 6 weeks. | Pilot randomized control trial | 38 cancer survivors | 15-minute recording, no positioning information mentioned; ECG device; HRV coherence ratio reported as primary | Relative to controls, the intervention group experienced trends toward improvements in stress, distress, fatigue, PTSD, and depression. HRV coherence ratio increased following biofeedback training in the intervention group while no change or decrease in coherence ratio occurred with control group. |
| Tsai (2021) | Taiwan | To assess what factors may influence the outcome of cancer related fatigue among female breast cancer patients undergoing chemotherapy | Cohort study | 109 patients undergoing cancer treatment | 15-minute recording, no positioning information given; ECG device; frequency domain as primary | Anxiety, depression, and symptom distress contributed to cancer related fatigue with symptom distress, anxiety and sleep deprivation consistently high amongst those receiving chemotherapy. No predictive relationship was found between LF/HF ratio and fatigue levels. |
| Cheng (2019) | Taiwan | To assess if 12 weeks of mindfulness or Qigong training change outcomes for fatigue and HRV in cancer survivors | Randomized control trial | 66 cancer survivors | 5-minute recording; no positioning information given; PPG device; time domain as primary | Reduced cancer-related fatigue was present in both the mindfulness group and the Qigong group. SDNN and TP values also increased from baseline. |
| Riesenberg (2009) | Germany | To assess if intensive supervised aerobic exercise is efficacious in improving HRV, quality of life, and functional status for patients with lung cancer over a 28-day period in inpatient rehab? | Cohort study | 45 patients undergoing cancer treatment | 12-minute recording laying down; PPG device; time domain as primary | Exercise capacity and functional status (quality of life, fatigue) improved during rehabilitation. RMSSD increased post intervention. |
| Chuang (2010) | Taiwan | To assess if music therapy is effective in lowering anxiety, improving mood, and improving HRV when applied for a 2-hour session one time intervention in cancer survivors | Quasi-experimental | 23 cancer survivors | 5-minute recording, no positioning information given; ECG device; frequency domain as primary | Fatigue levels were lower after music therapy. Increase in HF power and decrease in LF/HF ratio and LF power were observed post intervention. |
| Fernandez-Lao (2011) | Spain | To investigate the immediate effects of myofascial release on HRV and mood state and to assess the attitude towards massage in breast cancer survivors suffering cancer-related fatigue | Crossover randomized control trial | 20 cancer survivors | 5min ECG, no positioning information given; ECG device; time domain as primary | Massage leads to an immediate increase in SDNN, RMSSD and HF and an improvement in mood. The impact of massage on cancer-related fatigue is modulated by the attitude of the patient towards massage, however no interaction was reported between belief and SDNN, RMSSD or HF values. |
| Chen (2019) | Taiwan | To assess the effect of a single music therapy intervention on multiple mood categories and HRV for women actively being treated for breast cancer. | Randomized control trial | 100 patients undergoing cancer treatment | 5-minute recording, no positioning information given; ECG device; frequency domain as primary | Women who completed single session music intervention reduced symptoms post intervention compared to control group. Higher LF/HF ratio reported at baseline, but no post intervention data was given. |
| Aoki (2023) | Japan | To assess if HRV would be useful in evaluating fatigue and quality of life of women with breast cancer undergoing radiotherapy. | Cohort study | 75 patients undergoing cancer treatment | 2-minute recording, sitting; PPG device; frequency domain as primary | Cognitive and physical fatigue were highest at baseline and decreased by midpoint of treatment. Similarly, a decrease in LF/HF ratio was seen by midpoint of treatment. |
| Masel (2016) | Austria | To assess if HRV a valid representation for subjective pain for patients in palliative care with cancer break through pain. | Pilot study | 10 patients undergoing cancer treatment | 24-hour recording, no positioning data given; ECG device; frequency domain as primary | Pain reduction was seen post opioid administration. Log LF/HF ratio decreased in patients with >2 point reduction on the NRS. |
| Lee (2022) | Taiwan | To assess if there is a difference in HRV measures and quality of life measures when given either Qigong, mindfulness intervention or no intervention in cancer survivors at least 1 month out of treatment. | Quasi-experimental | 125 cancer survivors | 5-minute recording, sitting; PPG device; time domain as primary | Both Qigong and mindfulness interventions had short-term effects in improving overall physical and mental health, and quality of life. Increase in SDNN and total power were noted in the short term, but decreased LF was reported in the long term. |
| Uchida (2017) | Japan | To assess if HRV is predictive of PACU emergent pain within a 12h hour window in post operative breast cancer surgery patients. | Retrospective study | 20 patients undergoing cancer treatment | No positioning or recording time information given; PPG device; frequency domain as primary | There was no significant difference in pain intensity between the two groups immediately after admission to PACU. Patients in the intervention group required no analgesic agents for at least 12 hours after surgery. Increased HF and decreased LF/HF ratio values were associated with adequate pain intervention. |
| Fagundes (2011) | USA | To assess if fatigue level is related to HRV or epinephrine at baseline and during/after a social stressor in breast cancer survivors. | Cohort study | 109 cancer survivors | Continuous recording, sitting; PPG device; time domain as primary | Higher fatigue was associated with decreased RMSSD values pre and post stressor. |
| Niederer (2013) | Germany | To assess if exercise over a 16-week period improves HRV measures and cancer related fatigue in both cancer survivors and patients undergoing cancer treatment. | Quasi-experimental | 45 cancer survivors and those undergoing treatment | 5-minute recordings, supine; PPG device; frequency domain as primary | Exercise programming improves quality of life and increases total power values. An increase in HF and decrease in LF was also observed post intervention. |
| Yesil (2018) | Turkey | To assess whether neuropathic pain is associated with changes in cardiac sympathovagal activity in patients with breast cancer. | Cohort study | 70 patients undergoing cancer treatment | 24-hour recording, no positioning information given; ECG device; time domain as primary | SDNN was lower and LF/HF ratio was higher in patients with neuropathic pain than in those without. |
| Fournié (2021) | France | To evaluate the feasibility of a 12-week program combining heart rate variability biofeedback and physical | Cohort study | 20 patients undergoing cancer treatment | 5-minute recording, seated with knees at 90°, feet flat on the floor, hands on | No changes in fatigue outcome measures noted despite a trend towards decreasing general/physical fatigue. SDNN and RMSSD did not change |
|  |  | exercise in hematologic patients. |  |  | thighs, palms facing upward, and eyes closed; PPG device; time domain as primary | over time, but increased LF and HRV coherence ratio were noted. |
| Murofushi (2023) | Japan | To assess if HRV can provide an objective measure for bone metastasis pain during a period of radiotherapy. | Pilot study | 11 patients undergoing cancer treatment | 24-hour recording, no positioning information given; PPG device; frequency domain as primary | For all samples, the NRS score showed no significant relationship with mean LF/HF ratio and HF. In patients with an NRS $\geq 4$, the NRS score was significantly associated with LF/HF ratio but not HF. |
| Guimond (2018) | Canada | To assess if measures of emotional regulation are associated with measures of psychological symptoms in patients with breast cancer receiving radiotherapy. | Cross-sectional | 81 patients undergoing cancer treatment | 5-minute recording, no positioning information given; PPG device; time domain as primary | Low emotional regulation was marginally associated with increased depression and fatigue between timepoints. HF was not associated with symptom change. |
| Uchida (2017) | Japan | To assess if low dose Remifentanyl effects post operative pain and HRV in breast cancer surgery patients. | Retrospective study | 20 patients undergoing cancer treatment | No positioning or recording time information given; PPG device; frequency domain as primary | After Remifentanyl administration, the value of the NRS decreased. There were no significant differences in HF and LF/HF ratio was found between groups. |

PPG= photoplethysmogram; SDNN= standard deviation of the normal-to-normal interval; TP= total power; HF= high frequency power; PTSD= post-traumatic stress syndrome; HRV= heart rate variability; ECG=electrocardiogram; LF/HF ratio= low frequency/high frequency ratio; RMSSD = square root of mean squared differences of successive normal-to-normal intervals; NRS= numeric rating scale; LF= low frequency; PACU= post anesthesia care unit

| **First author (year)** | **Country** | **Aim of study** | **Design** | **Participants recruited** | **Heart rate variability measurement details** | **Key relevant findings** |
| --- | --- | --- | --- | --- | --- | --- |
| Lee (2018) | Taiwan | To assess the effects of 12 weeks of Qigong exercise or stress management training on fatigue, fear of recurrence, quality of life, and heart rate variability in people who completed cancer treatment at least 1 month prior. | Randomized control trial | 99 cancer survivors | 5-minute recording seated or lying prone; PPG device; time domain as primary | After 12 weeks of both Qigong and stress management training, fatigue was reduced and increases in SDNN, TP and HF were observed. |
| Burch (2020) | USA | To assess if heart rate variability biofeedback treatment among cancer survivors can increase HRV coherence and reduce symptoms of pain, stress, fatigue, depression, distress, PTSD, and insomnia relative to control patients in 6 weeks. | Pilot randomized control trial | 38 cancer survivors | 15-minute recording, no positioning information mentioned; ECG device; HRV coherence ratio reported as primary | Relative to controls, the intervention group experienced trends toward improvements in stress, distress, fatigue, PTSD, and depression. HRV coherence ratio increased following biofeedback training in the intervention group while no change or decrease in coherence ratio occurred with control group. |
| Tsai (2021) | Taiwan | To assess what factors may influence the outcome of cancer related fatigue among female breast | Cohort study | 109 patients undergoing cancer treatment | 15-minute recording, no positioning information given; ECG device; | Anxiety, depression, and symptom distress contributed to cancer related fatigue with symptom distress, anxiety |
|  |  | cancer patients undergoing chemotherapy |  |  | frequency domain as primary | and sleep deprivation consistently high amongst those receiving chemotherapy. No predictive relationship was found between LF/HF ratio and fatigue levels. |
| Cheng (2019) | Taiwan | To assess if 12 weeks of mindfulness or Qigong training change outcomes for fatigue and HRV in cancer survivors | Randomized control trial | 66 cancer survivors | 5-minute recording; no positioning information given; PPG device; time domain as primary | Reduced cancer-related fatigue was present in both the mindfulness group and the Qigong group. SDNN and TP values also increased from baseline. |
| Riesenberg (2009) | Germany | To assess if intensive supervised aerobic exercise is efficacious in improving HRV, quality of life, and functional status for patients with lung cancer over a 28-day period in inpatient rehab? | Cohort study | 45 patients undergoing cancer treatment | 12-minute recording laying down; PPG device; time domain as primary | Exercise capacity and functional status (quality of life, fatigue) improved during rehabilitation. RMSSD increased post intervention. |
| Chuang (2010) | Taiwan | To assess if music therapy is effective in lowering anxiety, improving mood, and improving HRV when applied for a 2-hour session one time intervention in cancer survivors | Quasi-experimental | 23 cancer survivors | 5-minute recording, no positioning information given; ECG device; frequency domain as primary | Fatigue levels were lower after music therapy. Increase in HF power and decrease in LF/HF ratio and LF power were observed post intervention. |
| Fernandez-Lao (2011) | Spain | To investigate the immediate effects of myofascial release on HRV and mood state and to assess the attitude towards massage in breast cancer survivors suffering cancer-related fatigue | Crossover randomized control trial | 20 cancer survivors | 5min ECG, no positioning information given; ECG device; time domain as primary | Massage leads to an immediate increase in SDNN, RMSSD and HF and an improvement in mood. The impact of massage on cancer-related fatigue is modulated by the attitude of the patient towards massage, however no interaction was reported between belief and SDNN, RMSSD or HF values. |
| Chen (2019) | Taiwan | To assess the effect of a single music therapy intervention on multiple mood categories and HRV for women actively being treated for breast cancer. | Randomized control trial | 100 patients undergoing cancer treatment | 5-minute recording, no positioning information given; ECG device; frequency domain as primary | Women who completed single session music intervention reduced symptoms post intervention compared to control group. Higher LF/HF ratio reported at baseline, but no post intervention data was given. |
| Aoki (2023) | Japan | To assess if HRV would be useful in evaluating fatigue and quality of life of women with breast cancer undergoing radiotherapy. | Cohort study | 75 patients undergoing cancer treatment | 2-minute recording, sitting; PPG device; frequency domain as primary | Cognitive and physical fatigue were highest at baseline and decreased by midpoint of treatment. Similarly, a decrease in LF/HF ratio was seen by midpoint of treatment. |
| Masel (2016) | Austria | To assess if HRV a valid representation for subjective pain for patients in palliative care with cancer break through pain. | Pilot study | 10 patients undergoing cancer treatment | 24-hour recording, no positioning data given; ECG device; frequency domain as primary | Pain reduction was seen post opioid administration. Log LF/HF ratio decreased in patients with >2 point reduction on the NRS. |
| Lee (2022) | Taiwan | To assess if there is a difference in HRV measures and quality of life measures when given either Qigong, mindfulness intervention or no intervention in cancer survivors at least 1 month out of treatment. | Quasi-experimental | 125 cancer survivors | 5-minute recording, sitting; PPG device; time domain as primary | Both Qigong and mindfulness interventions had short-term effects in improving overall physical and mental health, and quality of life. Increase in SDNN and total power were noted in the short term, but decreased LF was reported in the long term. |
| Uchida (2017) | Japan | To assess if HRV is predictive of PACU emergent pain within a 12h hour window in post operative breast cancer surgery patients. | Retrospective study | 20 patients undergoing cancer treatment | No positioning or recording time information given; PPG device; frequency domain as primary | There was no significant difference in pain intensity between the two groups immediately after admission to PACU. Patients in the intervention group required no analgesic agents for at least 12 hours after surgery. Increased HF and decreased LF/HF ratio values were associated with adequate pain intervention. |
| Fagundes (2011) | USA | To assess if fatigue level is related to HRV or epinephrine at baseline and during/after a social stressor in breast cancer survivors. | Cohort study | 109 cancer survivors | Continuous recording, sitting; PPG device; time domain as primary | Higher fatigue was associated with decreased RMSSD values pre and post stressor. |
| Niederer (2013) | Germany | To assess if exercise over a 16-week period improves HRV measures and cancer related fatigue in both cancer survivors and patients undergoing cancer treatment. | Quasi-experimental | 45 cancer survivors and those undergoing treatment | 5-minute recordings, supine; PPG device; frequency domain as primary | Exercise programming improves quality of life and increases total power values. An increase in HF and decrease in LF was also observed post intervention. |
| Yesil (2018) | Turkey | To assess whether neuropathic pain is associated with changes in cardiac sympathovagal activity in patients with breast cancer. | Cohort study | 70 patients undergoing cancer treatment | 24-hour recording, no positioning information given; ECG device; time domain as primary | SDNN was lower and LF/HF ratio was higher in patients with neuropathic pain than in those without. |
| Fournié (2021) | France | To evaluate the feasibility of a 12-week program combining heart rate variability biofeedback and physical exercise in hematologic patients. | Cohort study | 20 patients undergoing cancer treatment | 5-minute recording, seated with knees at 90°, feet flat on the floor, hands on thighs, palms facing upward, and eyes closed; PPG device; time domain as primary | No changes in fatigue outcome measures noted despite a trend towards decreasing general/physical fatigue. SDNN and RMSSD did not change over time, but increased LF and HRV coherence ratio were noted. |
| Murofushi (2023) | Japan | To assess if HRV can provide an objective measure for bone metastasis pain during a period of radiotherapy. | Pilot study | 11 patients undergoing cancer treatment | 24-hour recording, no positioning information given; PPG device; frequency domain as primary | For all samples, the NRS score showed no significant relationship with mean LF/HF ratio and HF. In patients with an NRS $\geq 4$, the NRS score was significantly associated with LF/HF ratio but not HF. |
| Guimond (2018) | Canada | To assess if measures of emotional regulation are associated with measures of psychological symptoms in patients with breast cancer receiving radiotherapy. | Cross-sectional | 81 patients undergoing cancer treatment | 5-minute recording, no positioning information given; PPG device; time domain as primary | Low emotional regulation was marginally associated with increased depression and fatigue between timepoints. HF was not associated with symptom change. |
| Uchida (2017) | Japan | To assess if low dose Remifentanyl effects post operative pain and HRV in breast cancer surgery patients. | Retrospective study | 20 patients undergoing cancer treatment | No positioning or recording time information given; PPG device; frequency domain as primary | After Remifentanyl administration, the value of the NRS decreased. There were no significant differences in HF and LF/HF ratio was found between groups. |

A majority of the participants across studies were middle aged (ranging from 48-65 years). All but 3 of the studies (34–36) reported a predominant female population, with 10 having an all-female sample (22–31). One study did not provide gender information (37). Studies took place in Taiwan (N= 6)(22, 24, 31, 33, 38, 39), Japan (N = 4)(25, 26, 30, 36), United States of America (USA) (N = 2)(27, 40), Germany (N = 2)(34, 37), Canada (N = 1)(29), France (N = 1)(35), Turkey (N = 1)(28), Spain (N = 1)(23), and Austria (N = 1)(32). Only 2 studies provided detailed data on race or ethnicity, and these studies took place in the USA (27, 40).

Fatigue was the most common symptom discussed (N = 14)(22–25, 27, 29, 31, 33–35, 37–39), followed by pain (N = 5)(26, 30, 32, 36). Although 1 study assessed neuropathic pain,(28) no studies were found on non-painful neuropathic symptoms, and only 1 study assessed multiple symptoms (pain and fatigue) (40). Four of the included studies were focused on the psychological effects of cancer (mood, anxiety, depression) as a primary measure, with fatigue as a secondary measure of interest (22, 23, 31, 39).

### Heart rate variability measures

All 19 studies utilized electrocardiogram (ECG) (N=6) or photoplethysmography (PPG) (N=13) equipment in the form of wrist watches, handheld or chest strap tools for HRV measurement. Ten of 19 studies conducted HRV measurements for 5 minutes or less.(23–25, 29, 31, 33–35, 38, 39) 3 of 19 reported between 12 and 15-minute recording times.(22, 37, 40) Furthermore, 4 of 19 reported continuous measurement for a maximum of 24 hours.(27, 28, 32, 36) Two studies did not report on the duration of the recording.(26, 30) Studies varied in participant positioning during HRV recording and in level of control for confounding factors (i.e. caffeine, physical activity), with 7 of 19 studies failing to include this information in the manuscript (22, 26, 30, 32, 33, 36, 39).

### Heart rate variability and fatigue

Fatigue was a primary outcome for 9 studies (24, 25, 27, 29, 33–35, 37, 38). Participants included cancer stages from stage I-IV, and sample sizes ranged from 17-125. Measures of fatigue were heterogeneous amongst the studies, and included the cancer fatigue scale (N= 3)(25, 33, 38), multidimensional fatigue inventory (MFI-20) (N=2)(35, 37), multidimensional fatigue symptom inventory (MFSI-SF) (N= 2)(24, 27), fatigue symptom inventory (N=1)(29) and the EORTC Core Quality of Life questionnaire (EORTC-QLQ C30) (N=1)(34).

Five of the 9 studies reported time domain measures. Standard deviation of normal-to-normal RR intervals (SDNN) was utilized for three studies (33, 35, 38). Root mean square of successive differences (RMSSD) was reported as a primary time domain variable in 2 studies (27, 37). Of these 5 studies, only one did not report secondary frequency domain variables (LF, HF, LF/HF ratio) (37). Furthermore, four of 9 studies only reported frequency domain variables (24, 25, 29, 34). All 9 studies reported longitudinal HRV measures at distinct time points (pre intervention, post intervention, follow up). Time periods between baseline and follow up measures were variable, with final follow up outcome assessment ranging between 12-28 weeks after initial intervention. HRV devices were worn for 5-15 minutes throughout the data collection process for all 9 studies, and 8 of the fatigue related studies reported controlling for confounding factors of HRV including quiet sitting, decreasing caffeine (24, 25, 27, 29, 34, 37, 38), and specific instructions like “relax and breathe normally”(35).

Across 4 of the studies (27, 33, 37, 38), a decrease in fatigue scores was associated with increased time domain HRV measures (SDNN, RMSSD). Only one study found no change in time domain measures amongst groups (35). The relationships between fatigue scores and frequency domain measures (LF/HF ratio, HF, LF, TP) were mixed with two studies reporting no change in the trials (29, 35), one study reporting decreased LF/HF ratio and fatigue at midpoint of treatment (25), and 3 studies reporting increased total power with decreased fatigue (33, 34, 38).

### Studies using heart rate variability with fatigue as secondary measures

Four studies included fatigue measures as a secondary focus, with a primary focus on emotional stressors like anxiety, depression or fear of cancer recurrence (22, 23, 31, 39). Like the studies with fatigue as a primary outcome, sample sizes varied from 20-100 participants (23, 31, 39). All 4 studies utilized ECG as a primary source of HRV measurement, with recordings lasting 5-15 minutes, with 2 reporting time domain variables as primary HRV outcomes (SDNN, RMSSD)(23, 39), and all four studies reported frequency domain measures (LF/HF ratio, TP, HF, LF). Three of the studies found that an improvement in HRV variables (increased SDNN, RMSSD, HF and decreased LF/HF ratio) occurred with decreased fatigue (23, 31, 39). Only one study found no predictive relationship between HRV and cancer related fatigue, and though this study had the largest sample size, it was also the only study involving participants currently undergoing cancer treatment (22).

Across these 13 studies, a decrease in fatigue scores typically occurred with an increase in time domain HRV measures, with only 1 of 7 studies that reported on time domain measures not finding this relationship. There was less consistency across fatigue related studies that used frequency domain HRV measures, although most also found a relationship. The GRADE for the evidence on HRV as a physiological marker for fatigue is **low** due to risk of bias (incomplete outcome data in 4 studies and lack of accounting for confounders in 2 studies).

### Heart rate variability and pain

Of the 4 studies with pain as the primary symptom under investigation, participants ranged in cancer stage of treatment from post operative to palliative care, and all of the studies had small sample sizes (n≤ 20). All 4 studies utilized the 0-10 numeric rating scale (NRS) for pain intensity, and none reported any additional pain variables. Additionally, all 4 studies utilized frequency domain HRV measures including high frequency (HF), low frequency (LF), total power (TP), and LF/HF ratio; and only one study reported a time domain variable (pNN50). Only 2 of the studies reported recording time frame (24 hours for both) (32, 36). HRV measurement with a PPG based monitoring system was closely tied to time periods of medication administration for pain management in 3 of 4 studies (26, 30, 32). Importantly, none of the four studies reported controlling for confounding factors pertaining to HRV measurement (e.g., time of day, phase of menstrual cycle, dietary and exercise considerations).

Three studies reported on acute pain changes (i.e., post operative and breakthrough pain)(26, 30, 32) and one on chronic pain associated with bony metastasis (36). Those studies on acute pain reported significant differences in frequency domain variables, mainly LF/HF ratio, with improving NRS scores (26, 30, 32). Two studies reported on the role of pain free post operative period in regulating HRV (LF/HF ratio and HF) (26, 30). With breakthrough pain specifically, an NRS reduction of greater than 2 points equated to a decrease in LF/HF ratio and improving function (32). With radiotherapy, higher NRS scores of greater than or equal to 4 were associated with LF/HF ratio changes with no significant association found with NRS scores below 4 (36).

Collectively, these studies suggest that moderate changes in pain intensity (NRS change of 2-4) may be associated with changes in HRV. However, the GRADE for this evidence is **very low** due to high risk of bias (incomplete reporting of data and lack of accounting for confounders) and imprecision (small sample sizes).

### Heart rate variability and neuropathy

Only one study mentioned neuropathy as a symptom of interest in conjunction with pain measures (28). Categorized as neuropathic pain, the study focused on breast cancer patients, and no cancer stage information was given. Unlike the other studies on pain, this study utilized the Leeds Assessment of Neuropathic Symptoms and Signs (LANSS) scale to differentiate between neuropathic and nociceptive pain descriptors, not intensity (28). ECG was utilized to measure HRV, with SDNN being the HRV variable of interest. Other time domain variables (SD of the mean of the RR intervals (SDAAN), SD of the NN interval (SDDN index) and frequency domain variables (total power (TP), LF/HF ratio) were also collected. Time domain variable values were found to be lower and frequency domain variable values were found to be higher in those with neuropathic pain (28). The GRADE for the evidence on HRV as a physiological marker for neuropathy is **very low** due to high risk of bias (lack of accounting for confounders) and imprecision (only 1 study).

## Discussion

HRV is a complex measure involving the fluctuation in the time intervals between heartbeats. These changes can influence regulatory outputs like blood pressure, heart and vascular tone, which influences adaptation to both physical and psychological stressors (41). This study aimed to assess the relationship of HRV to comorbid symptoms (i.e., pain, fatigue, and neuropathy) in cancer patients and survivors. Nineteen studies were identified that utilized HRV and assessed pain or fatigue. However, they varied in their purpose and application of HRV with some using HRV to determine the impact of symptoms on autonomic function and others using HRV as a measure of systemic response to a targeted intervention.

### HRV confounders

HRV measurements can be influenced by a variety of variables, including age, sex, and breathing rate (41). Numerous lifestyle factors including stress level, smoking, fitness level, caffeine intake and pharmacology, can also have an influence on HRV metrics (42). Thus, control and identification of these confounding factors is of upmost importance to decrease error in HRV interpretation. In our sample of studies, 7 of 19 included a description of controlling for potential confounders of HRV.

Additionally, sampling time can have an influence on accuracy of HRV metrics. The “gold standard” for clinical assessment is a 24 hour recording, as these recordings are shown to have greater predictive power (41). Short term measurements, defined as 5 minute measurements, or ultra short term measurements (<5 minutes) have been found to correlate poorly with the 24-hour indices (41). Of the 19 articles reviewed, 10 reported short term or ultra short-term measurements, and only 4 utilized 24-hour measurements. This may be a contributing factor to the heterogeneity in findings.

Further contributing to the heterogeneity, HRV can be divided into time, frequency, and spectral domain measures. Notably, only time and frequency domain measures were reported, as none of the 19 studies assessed spectral domain measures. Time domain HRV indices (i.e. SDNN, RMSSD, pNN50, HRmax-HRmin) quantify the amount of variability observed during a recording time frame (<1 minute to >24 hours) (41). The SDNN has been referred to as the “gold standard” HRV variable for medical stratification of cardiovascular risk when recorded over a 24-hour period (41, 43). In contrast, frequency domain HRV indices estimate the relative or absolute power over four frequency bands ultra-low frequency (ULF), very-low frequency (VLF), low-frequency (LF) and high-frequency (HF) (41). The ratio of LF to HF (LF/HF ratio) was originally interpreted as an estimate of sympathetic to parasympathetic balance, with low LF/HF ratio symbolizing parasympathetic dominance and high LF/HF ratio translating to sympathetic dominance (41). This interpretation contributes to the popularity of frequency domain variables being reported over time domain. However, the accuracy of this interpretation has been called into question due to the variable nature of sympathetic nervous system control. Moreover, LF/HF ratio, particularly short term measurements of LF/HF ratio, have been found to vary widely based on testing conditions (41). Kuss et al. (44) found that frequency domain variables showed up to a 10 fold increase in variation when compared to SDNN. Of the 19 studies examined, only 9 reported a time domain measure (SDNN or RMSSD), and 17 studies reported frequency domain measures, most commonly LF/HF ratio. The higher utilization of frequency domain variables over time domain variables goes against existing recommendations for HRV research, and it highlights the need for consensus guidelines on HRV research in cancer populations.

### Differences in Outcome Measures

The specific outcome measure chosen may also influence the observed relationships between HRV and pain or fatigue. The four studies on pain were noted to all utilize the same outcome measure (NRS), focusing on pain intensity with pharmacological interventions. However, pain is known to be multidimensional (45), and the pain NRS is a unidimensional measure. It is possible that changes in HRV are associated with aspects of pain other than intensity (e.g., pain interference on mood or sleep). In contrast, the studies that focused on fatigue utilized multidimensional measures of fatigue. Specifically, the Cancer Fatigue Scale (CFS), Fatigue Symptom Inventory (FSI) and Multidimensional Fatigue Symptom Inventory-Short Form (MFSI-SF) were all validated utilizing patients with a history of cancer and all measure physical, cognitive and emotional fatigue (46–48). However, outcome scores for these fatigue scales were usually reported as a total score change, with only one study reporting subscale changes (35). It is plausible that HRV is more closely associated with one aspect of fatigue than another (e.g., physical fatigue may be strongly correlated to HRV but emotional fatigue is not). Given the multidimensional nature of pain and fatigue, future studies on HRV should assess and report on multiple dimensions for the symptoms of interest.

### Limitations

Only articles in English were included, which is an important limitation of the review process used in this study. Additionally, our search yielded only one study on HRV and neuropathy, and this study focused on neuropathic pain. The lack of longitudinal studies on HRV and neuropathy may be influenced by the lack of consensus regarding the diagnostic criteria for chemotherapy induced peripheral neuropathy (CIPN)(8). Another potential reason for this may be that CIPN is only found with neurotoxic chemotherapies such as paclitaxel, cisplatin, and oxaliplatin which may not have been prescribed in a large portion of the included abstracts for review (49). Due to the heterogeneity of the HRV measurements and the relatively high risk of bias across studies, we did not conduct a meta-analysis as planned. Additionally, only 2 of 19 studies reported on race/ethnicity in their given populations, and 10 of 19 studies took place in Asia (Taiwan, Japan), which may limit the generalizability of these findings.

## Conclusion

HRV is growing in use as a physiological outcome measure, and the findings of this study suggest that it may be a useful tool to determine physiological load associated with fatigue and pain in cancer patients and survivors. Fatigue and pain are common comorbidities associated with antineoplastic treatment and while their physiological impact can be seen with HRV changes, the lack of consensus with HRV measurement can make its interpretation difficult. Large studies with low risk of bias are needed as the field continues to work toward better standardization of HRV protocols.

## Data Availability

All data produced in the present study are available upon reasonable request to the authors

## Acknowledgements

For peer reviewing the Medline (OVID) database search strategy, the authors would like to acknowledge the Research Services and Liaison Librarian David S. Gerstle, Ph.D., MS-ILS from the University of Toronto Mississauga. The authors would also like to acknowledge John Reynolds and Thilani Samarakoon for their input on methodology.

---

**Appendix 1.**
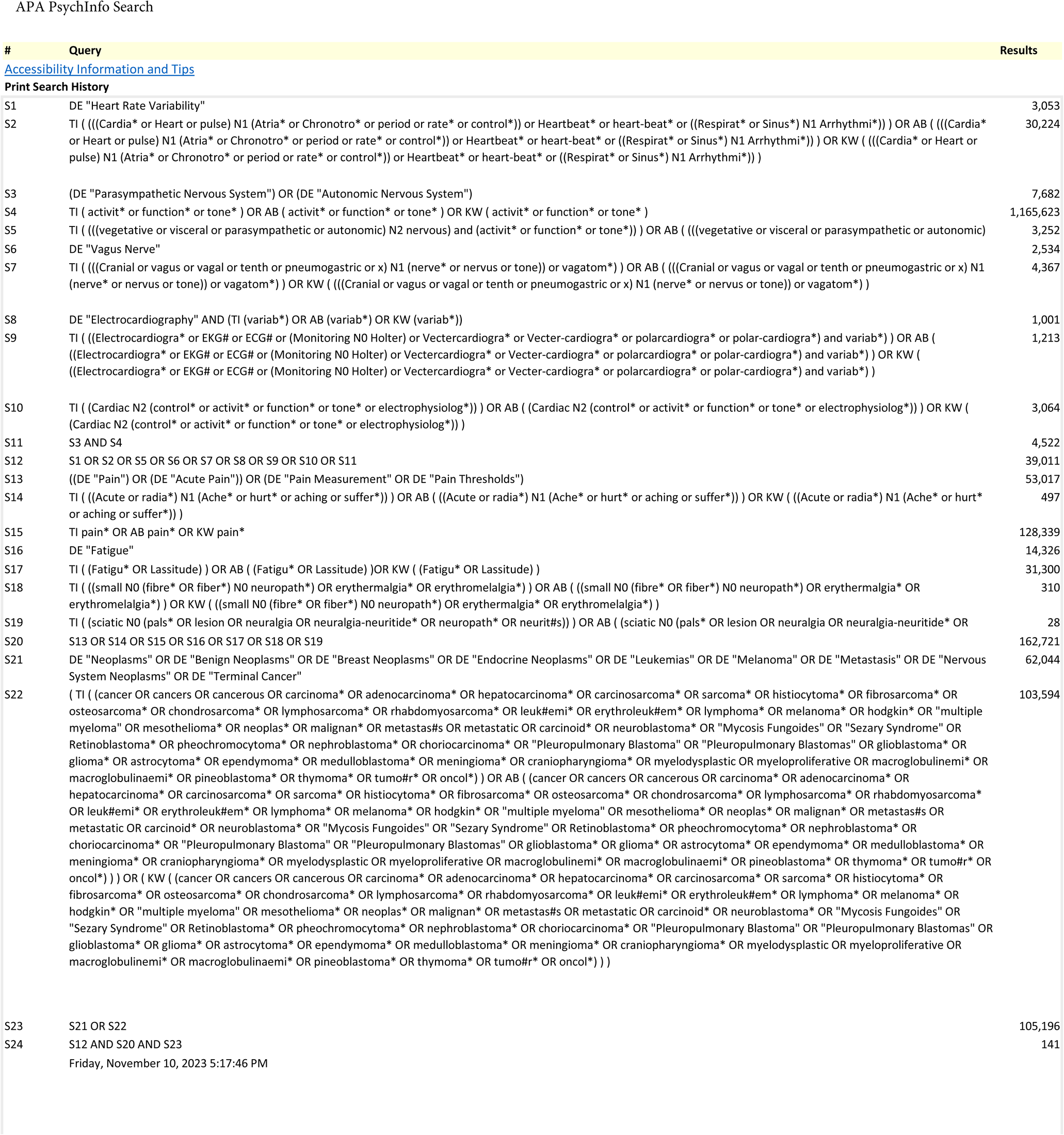
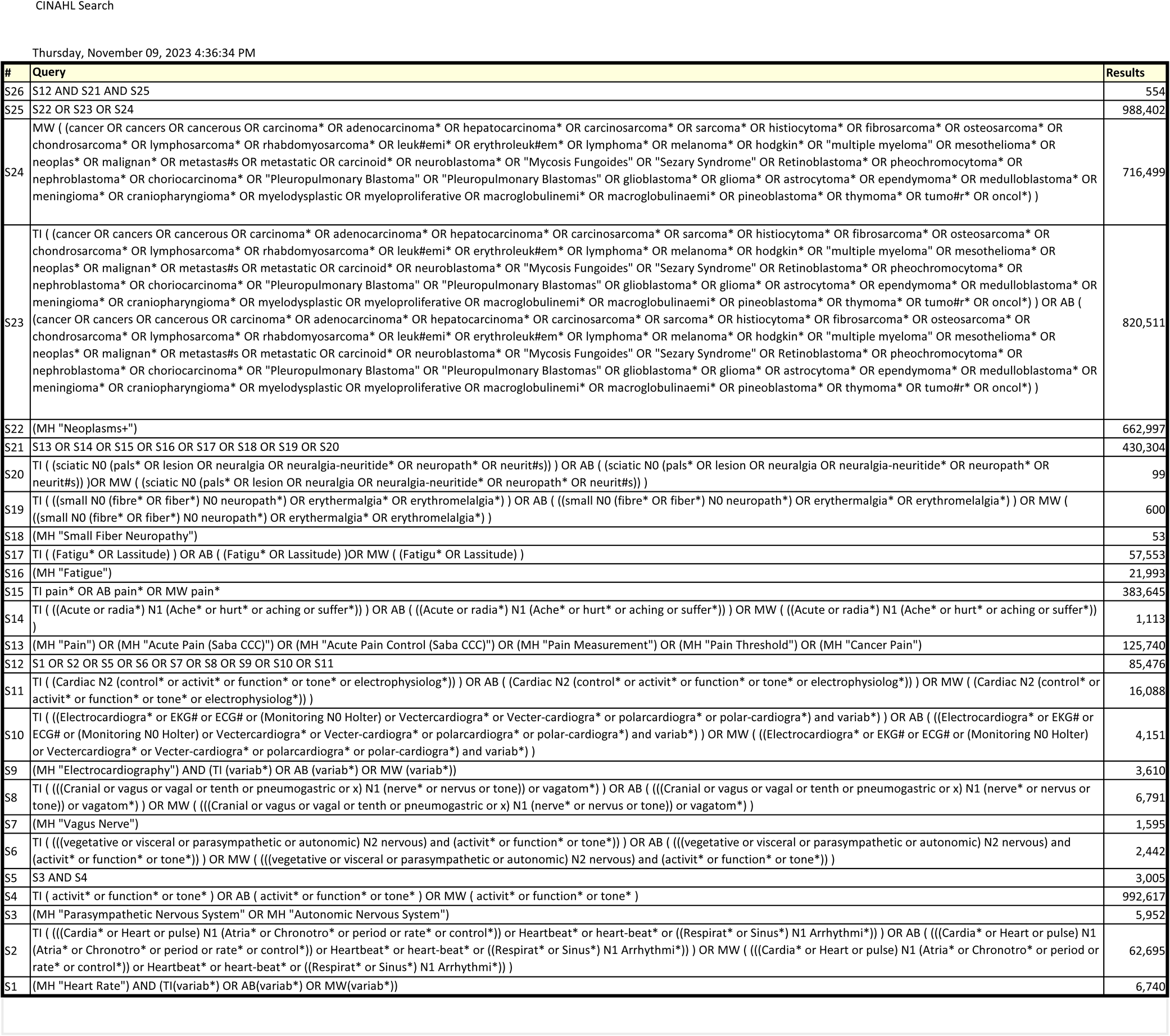
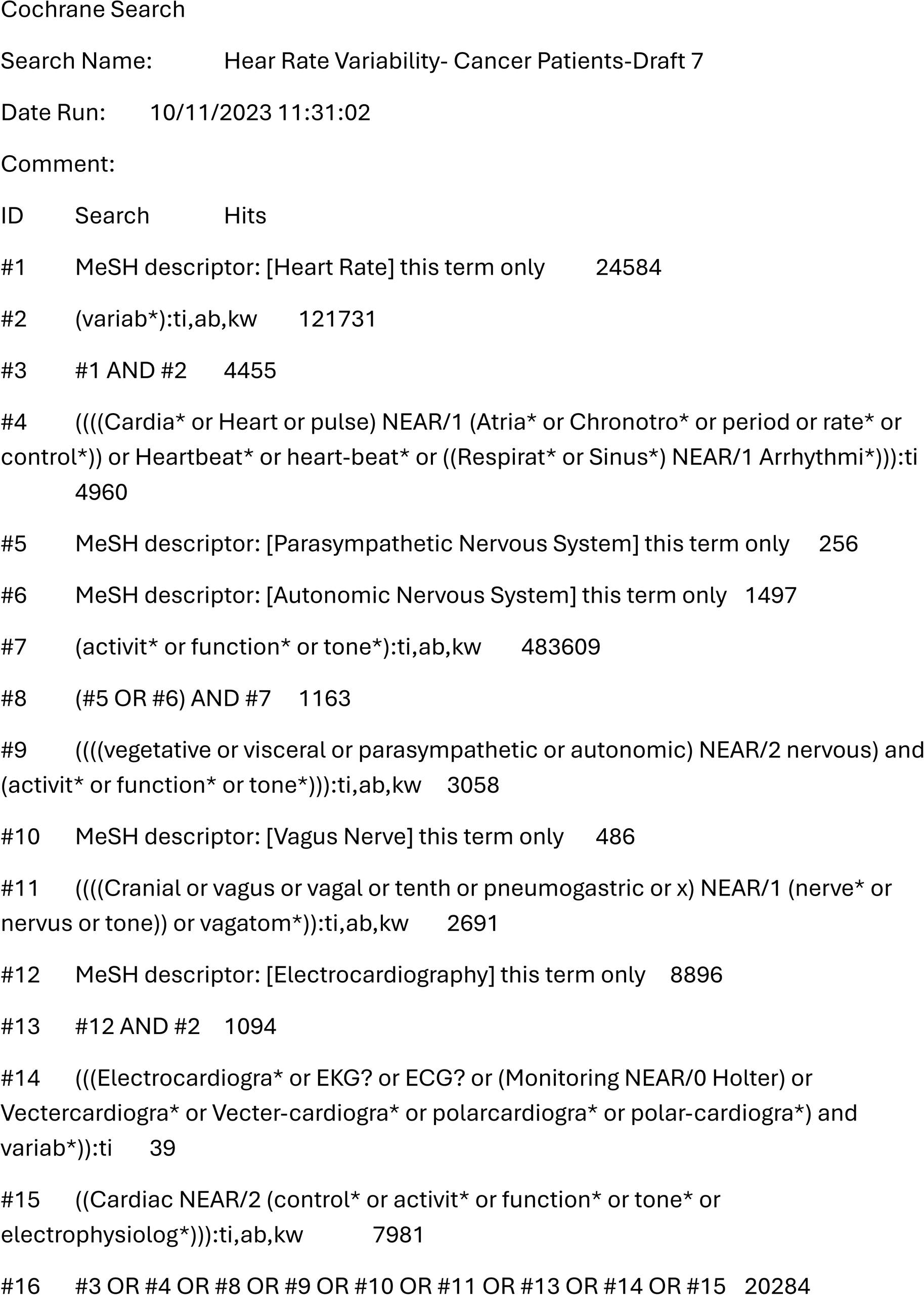
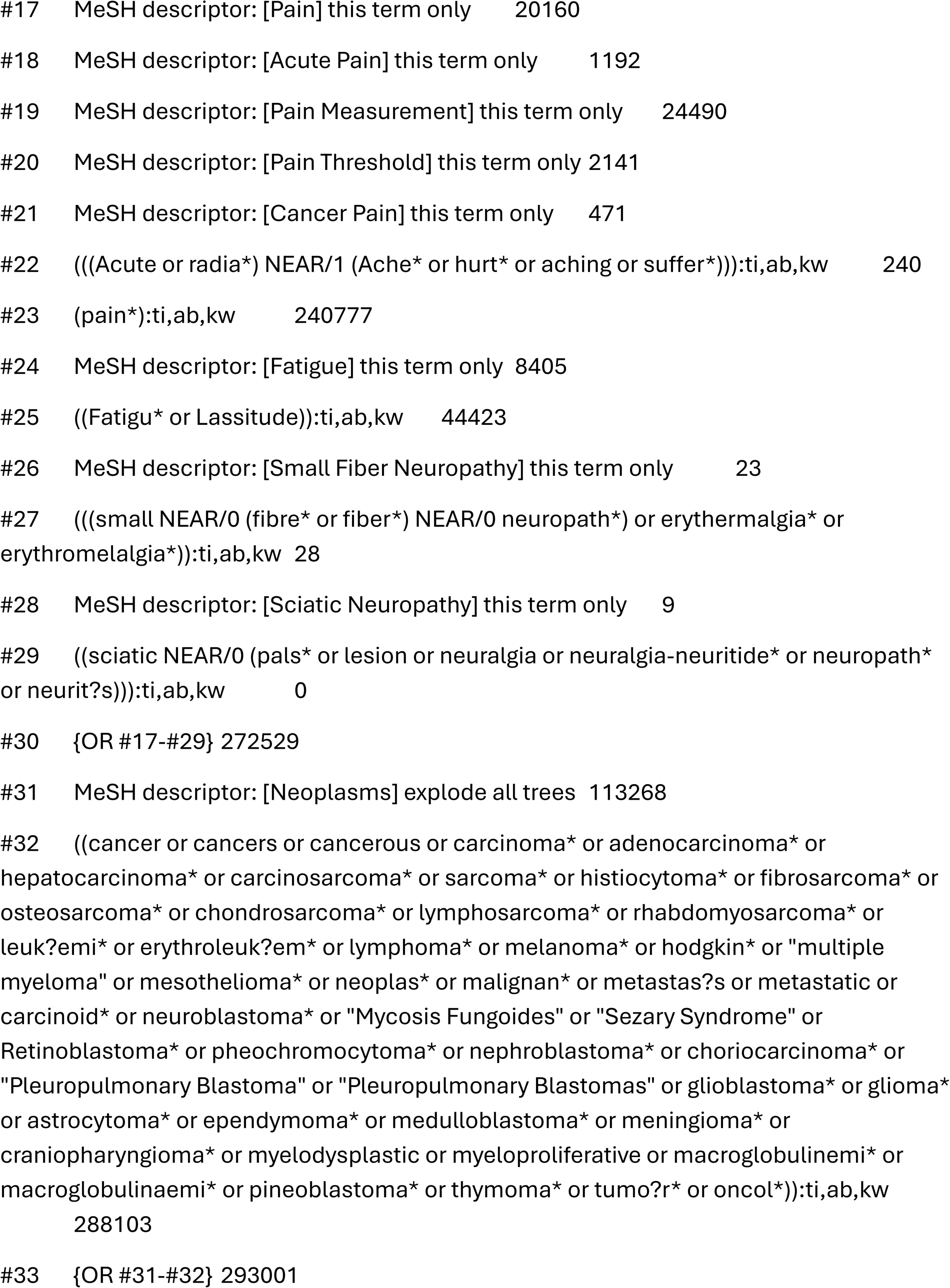
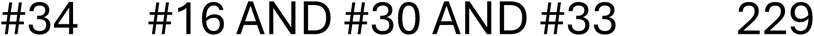
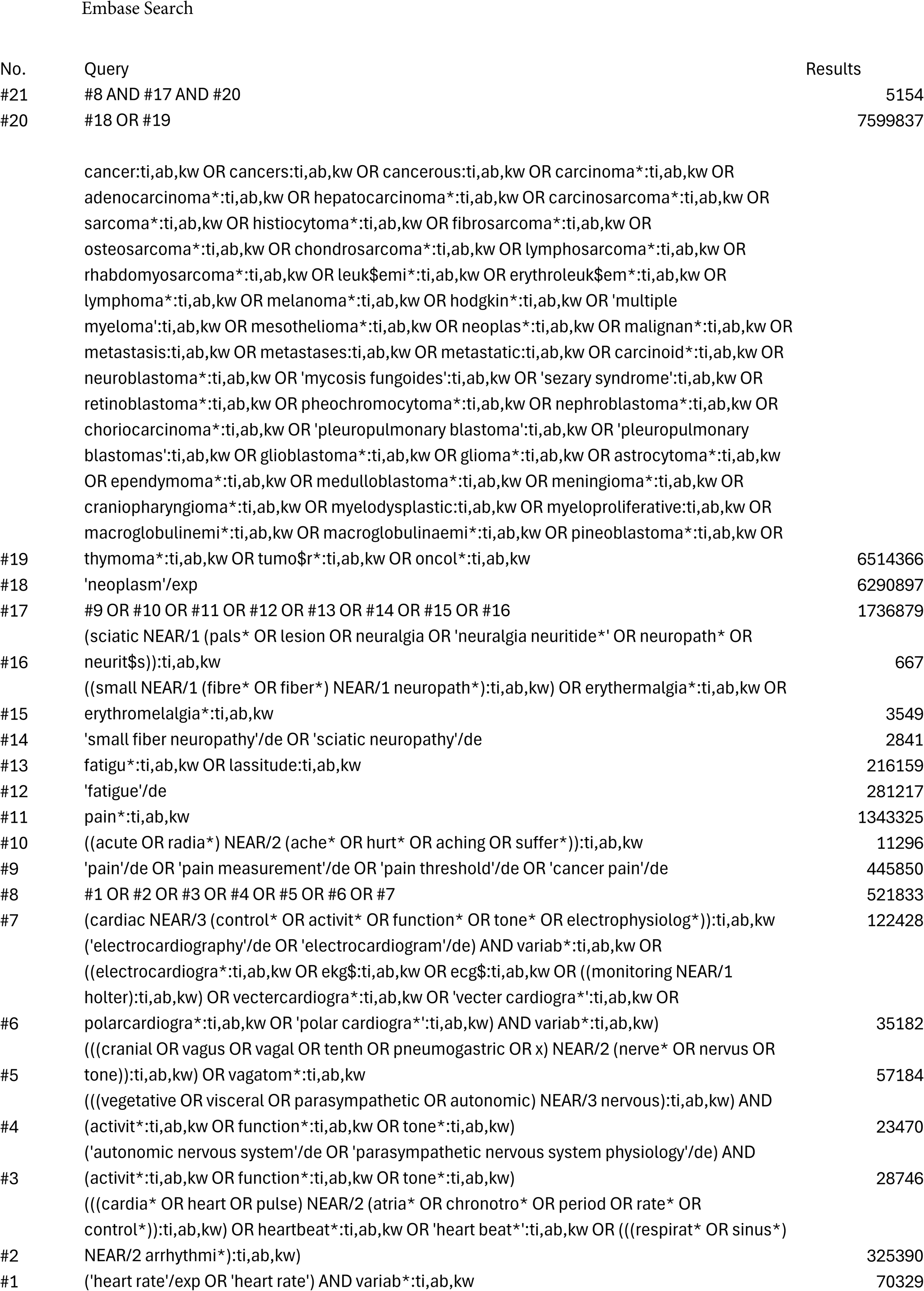
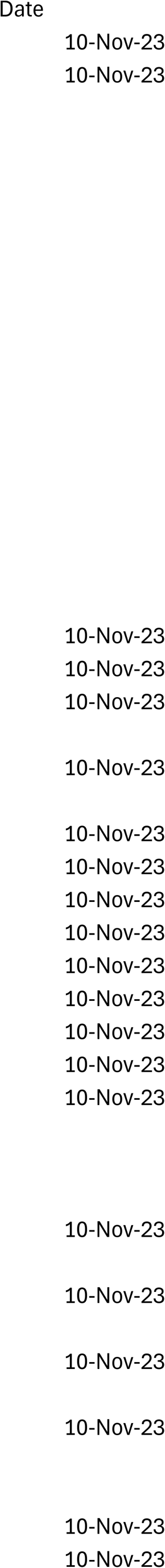
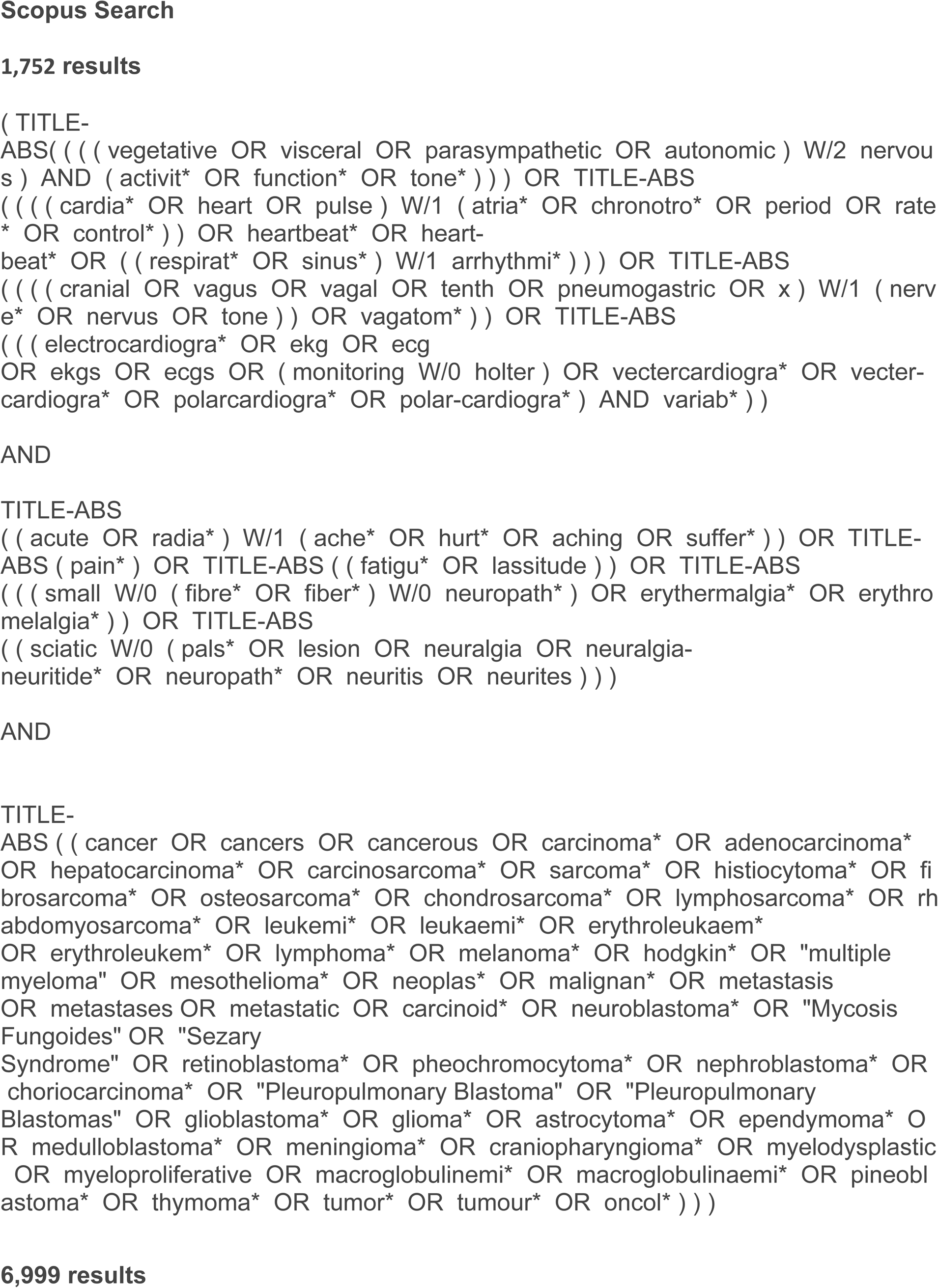
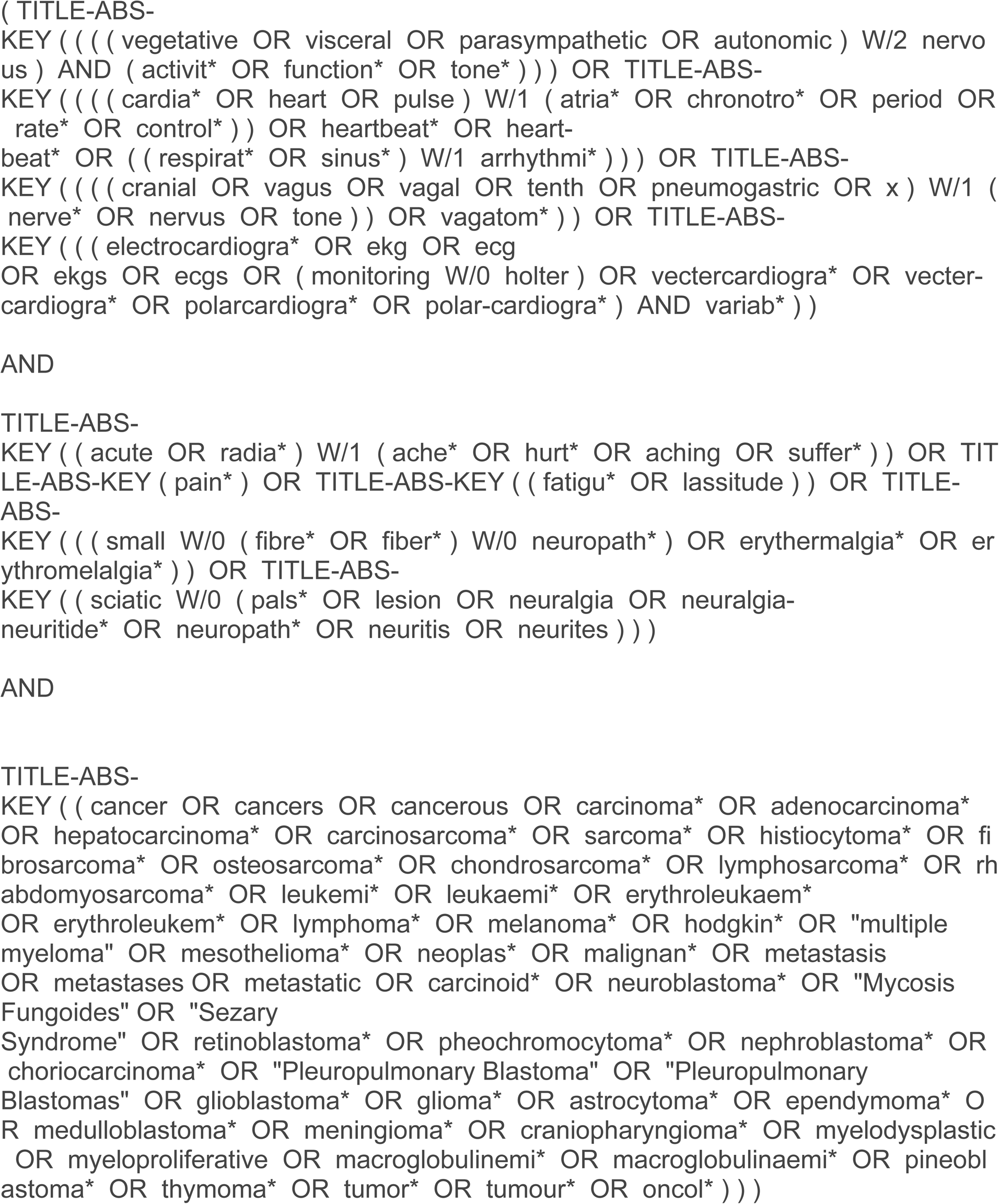
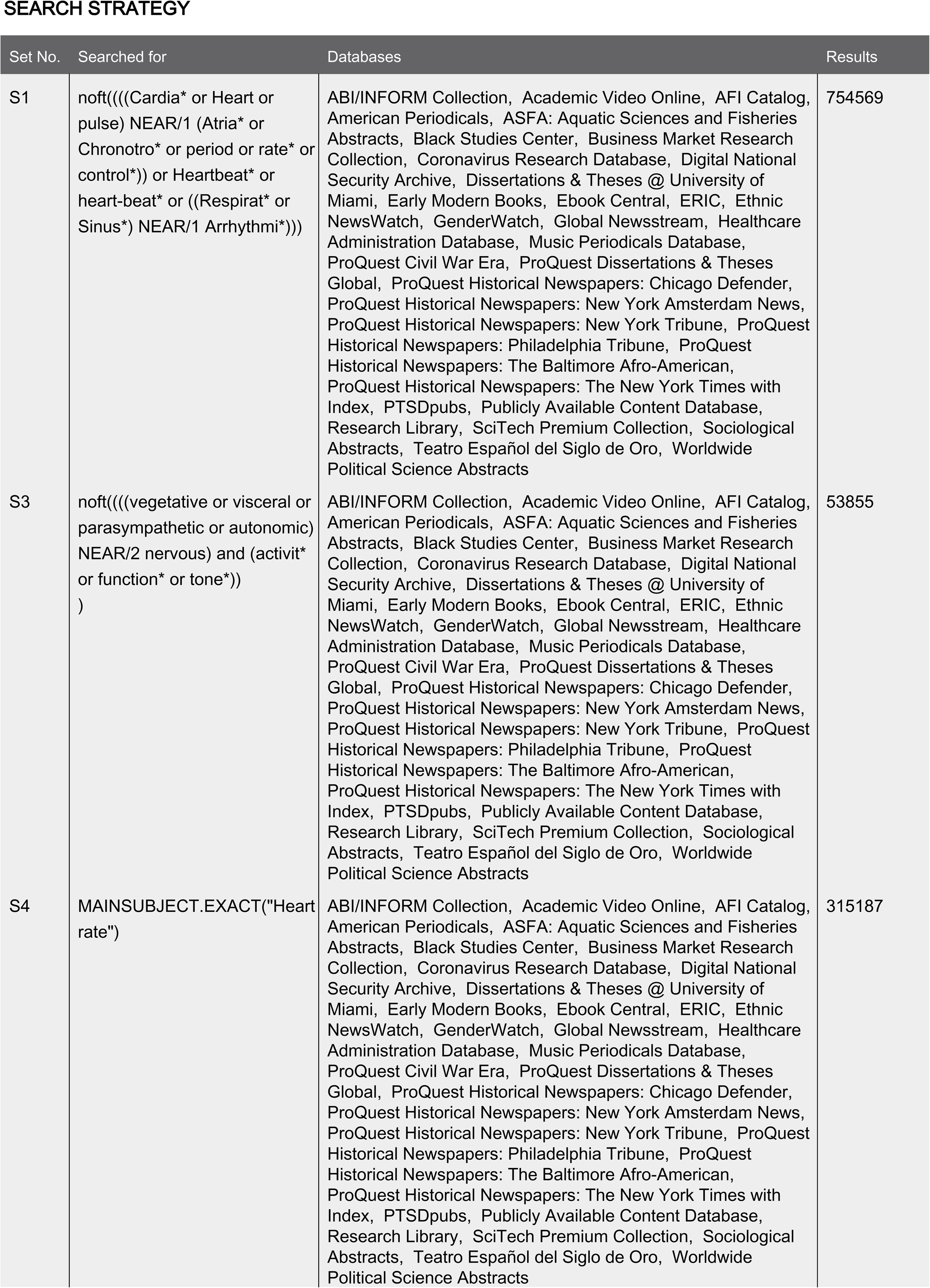
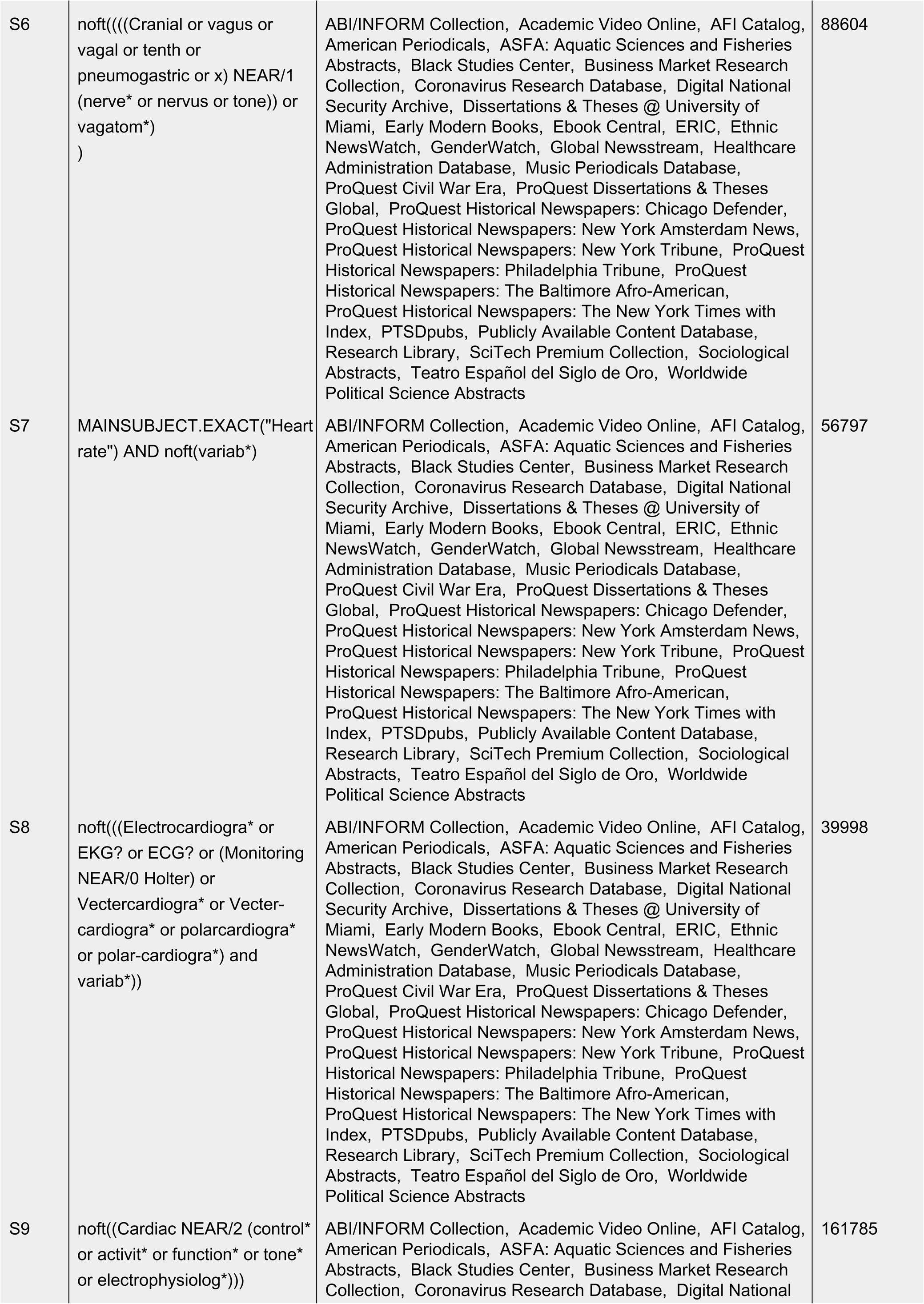
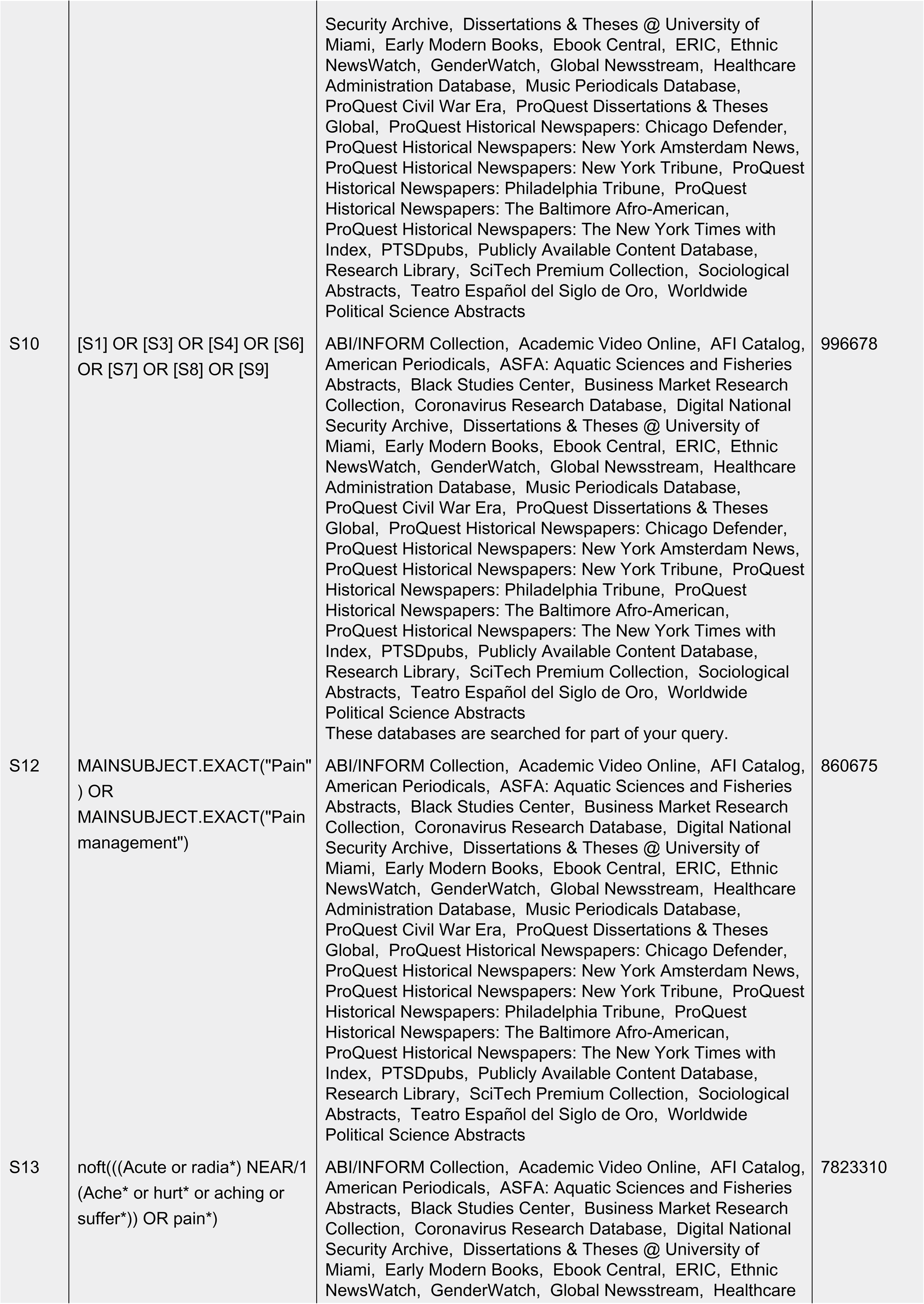
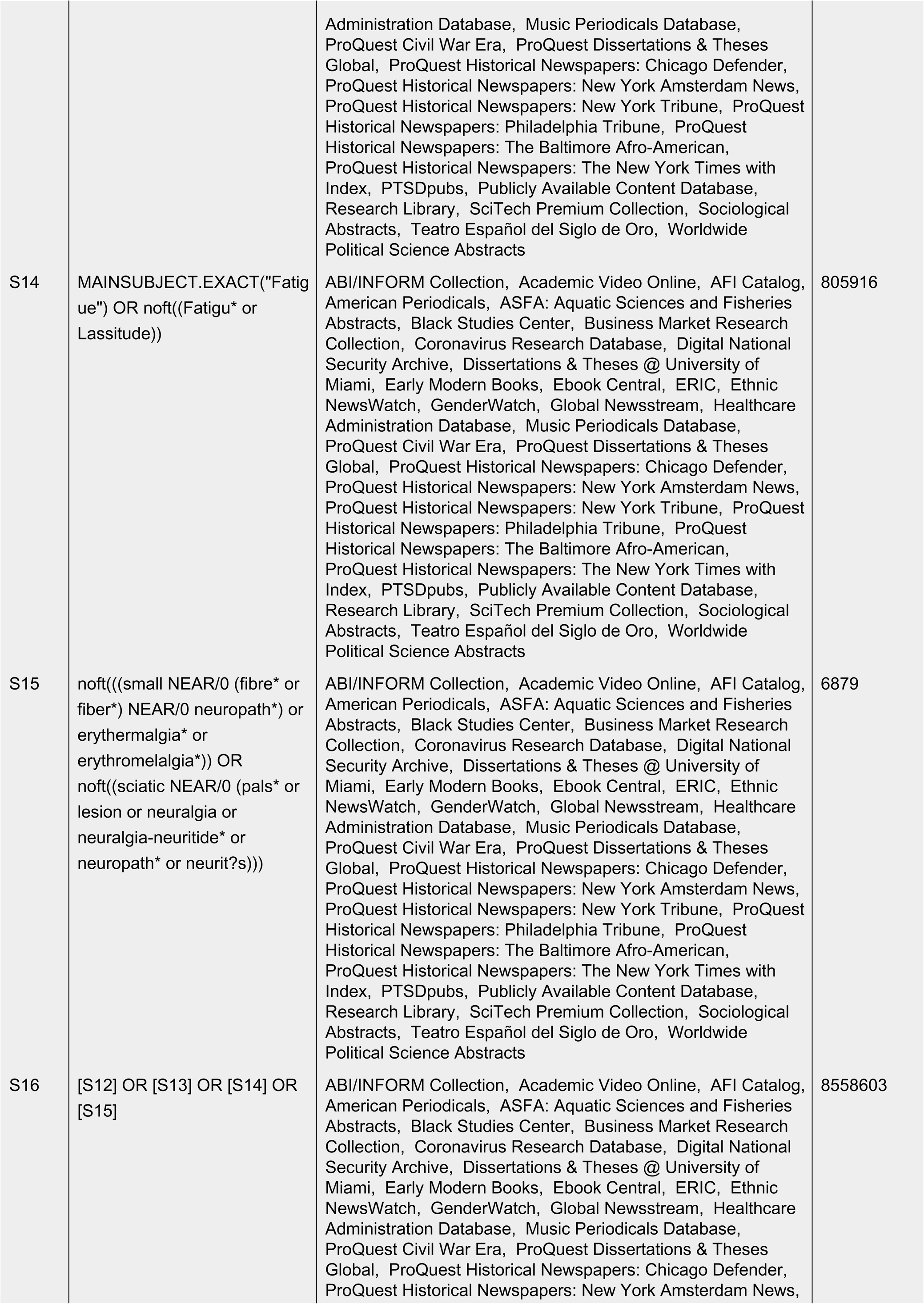
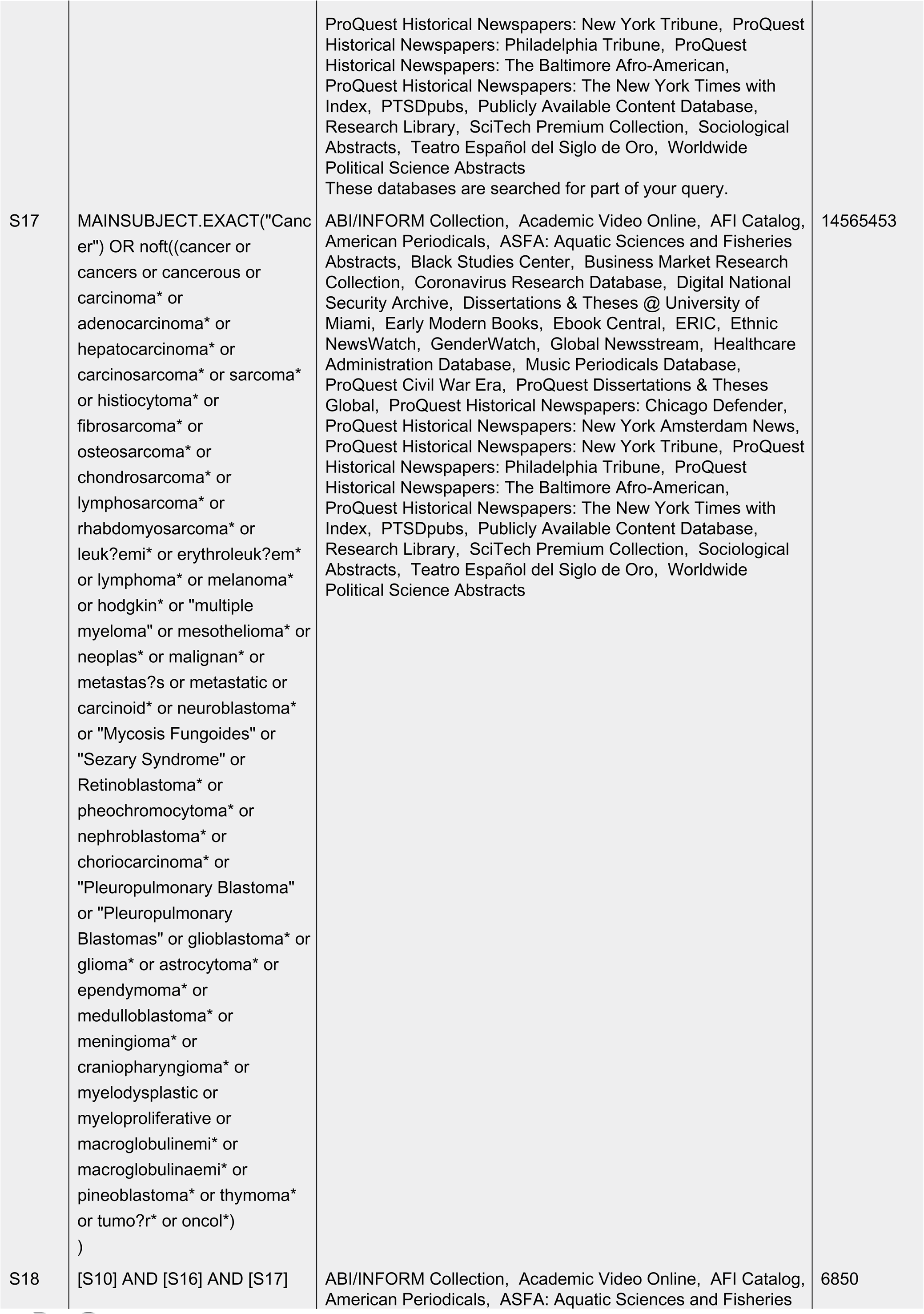
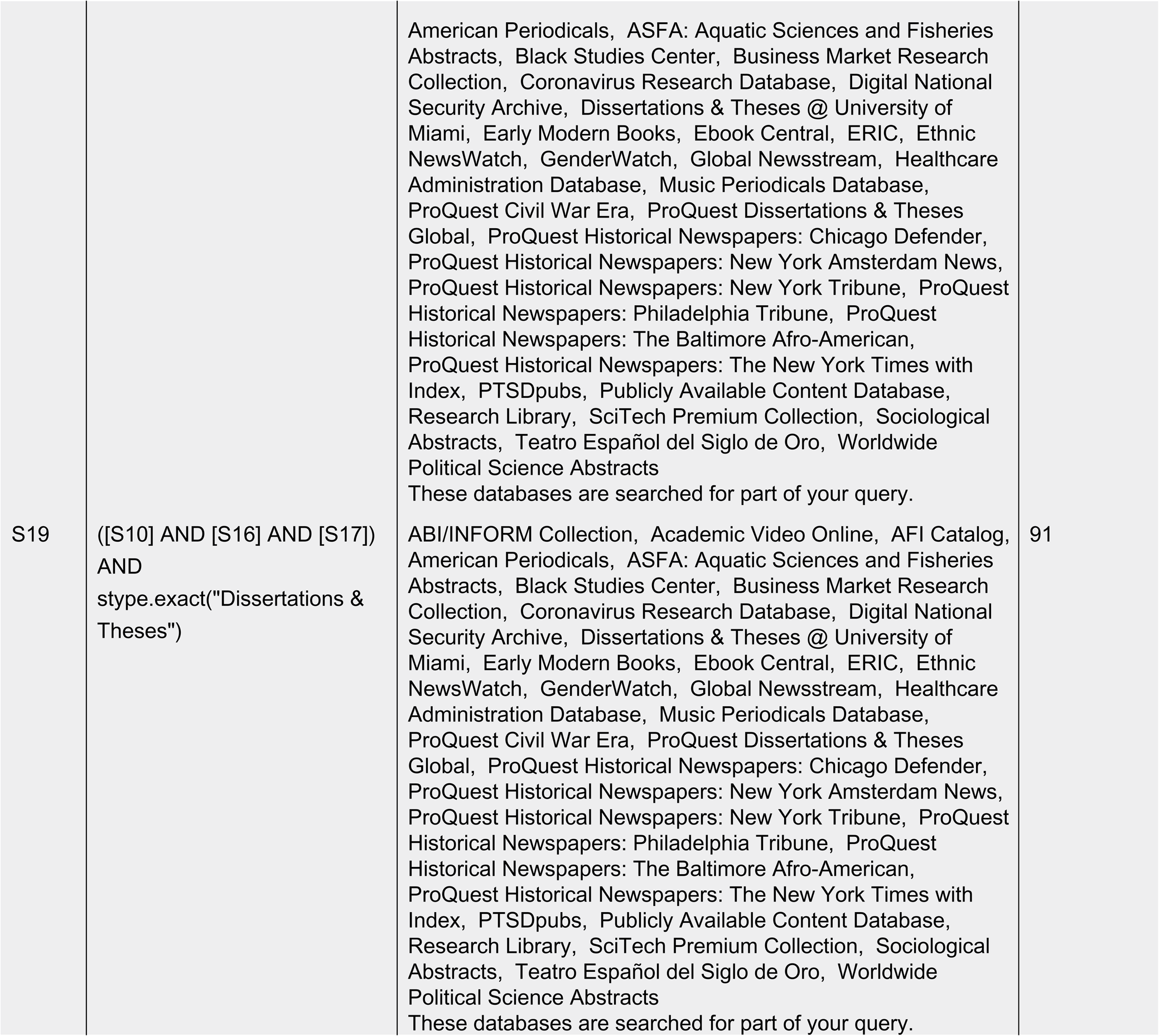
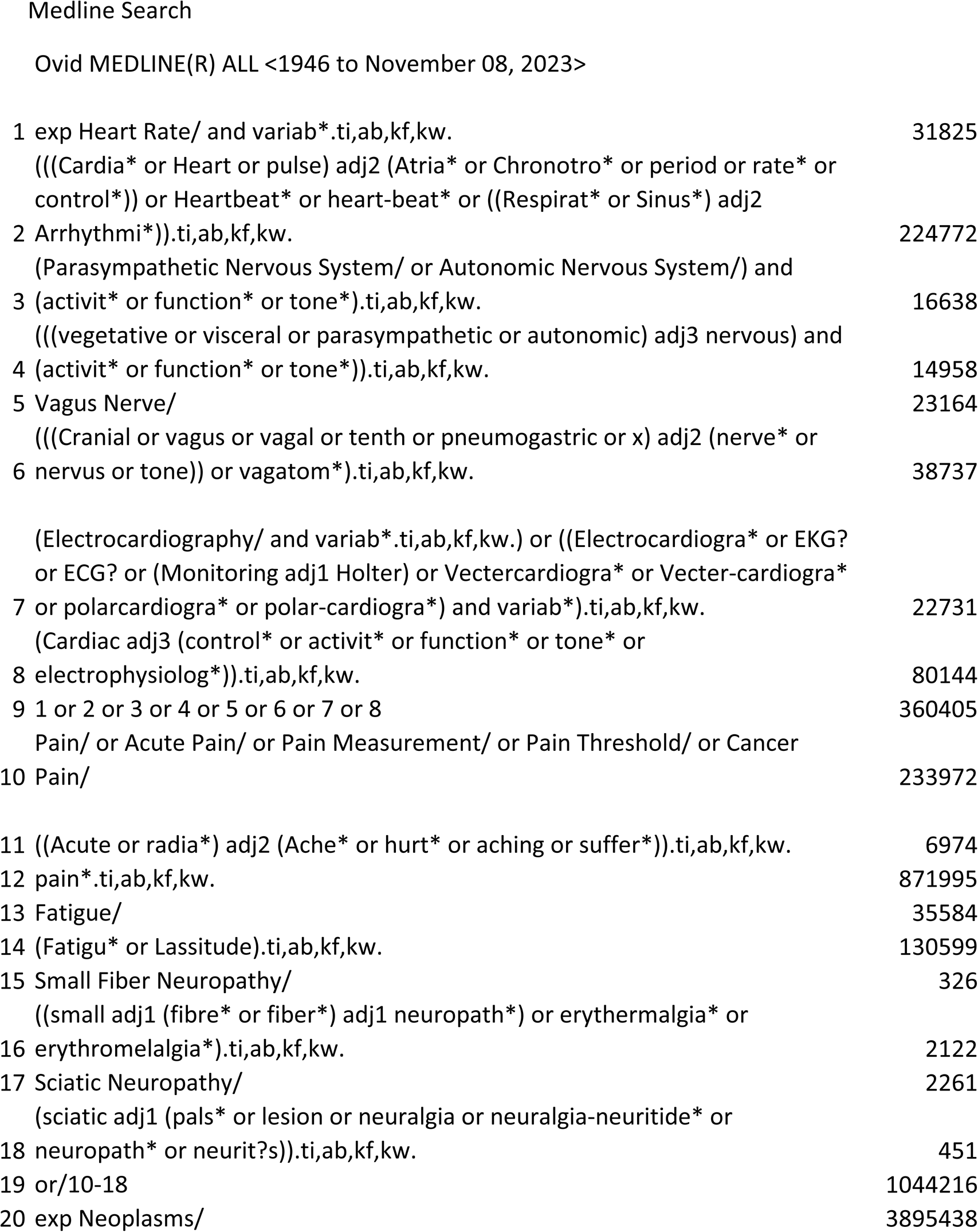
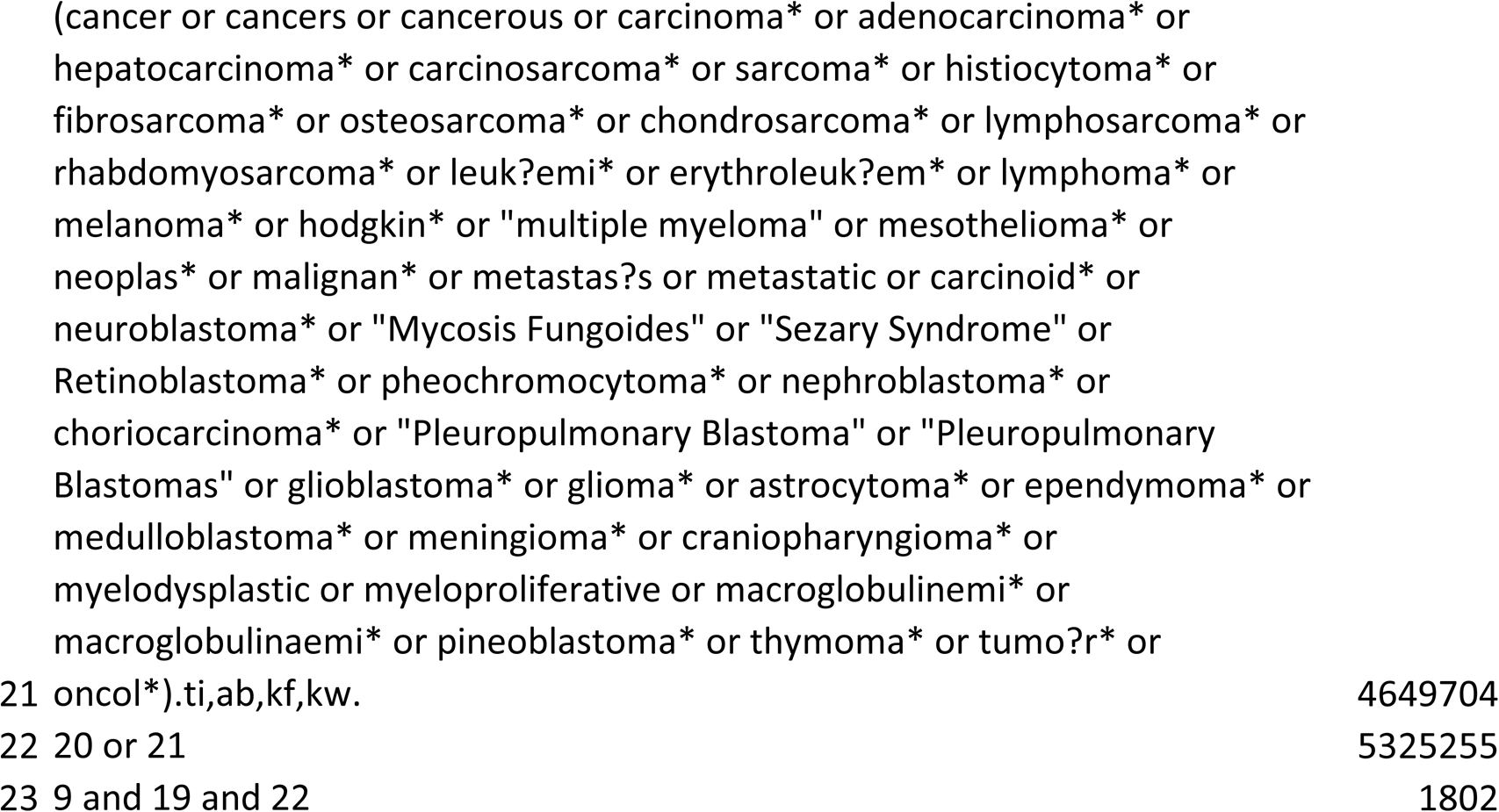
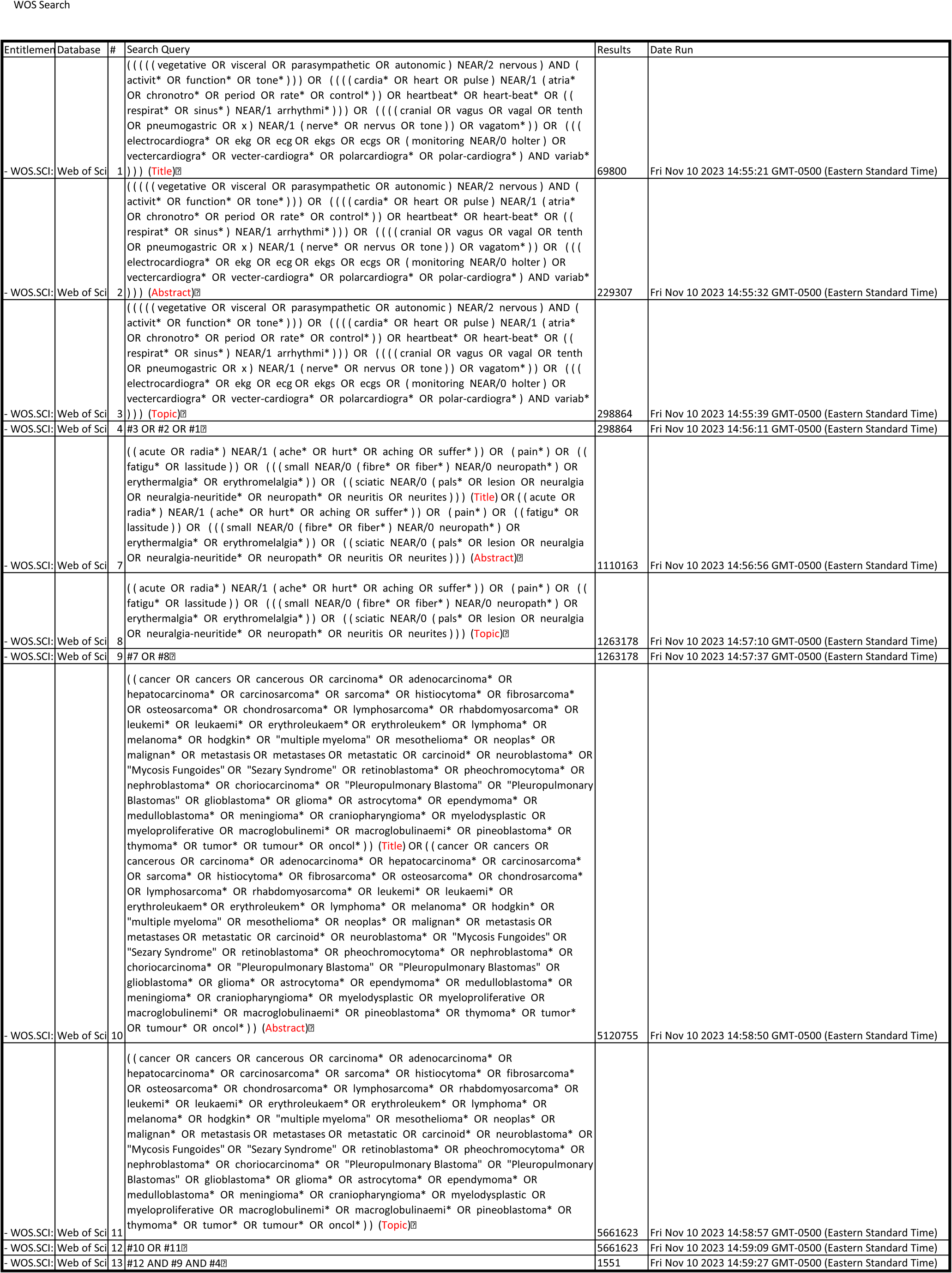
Full Search Strategy.

**Appendix 2.**
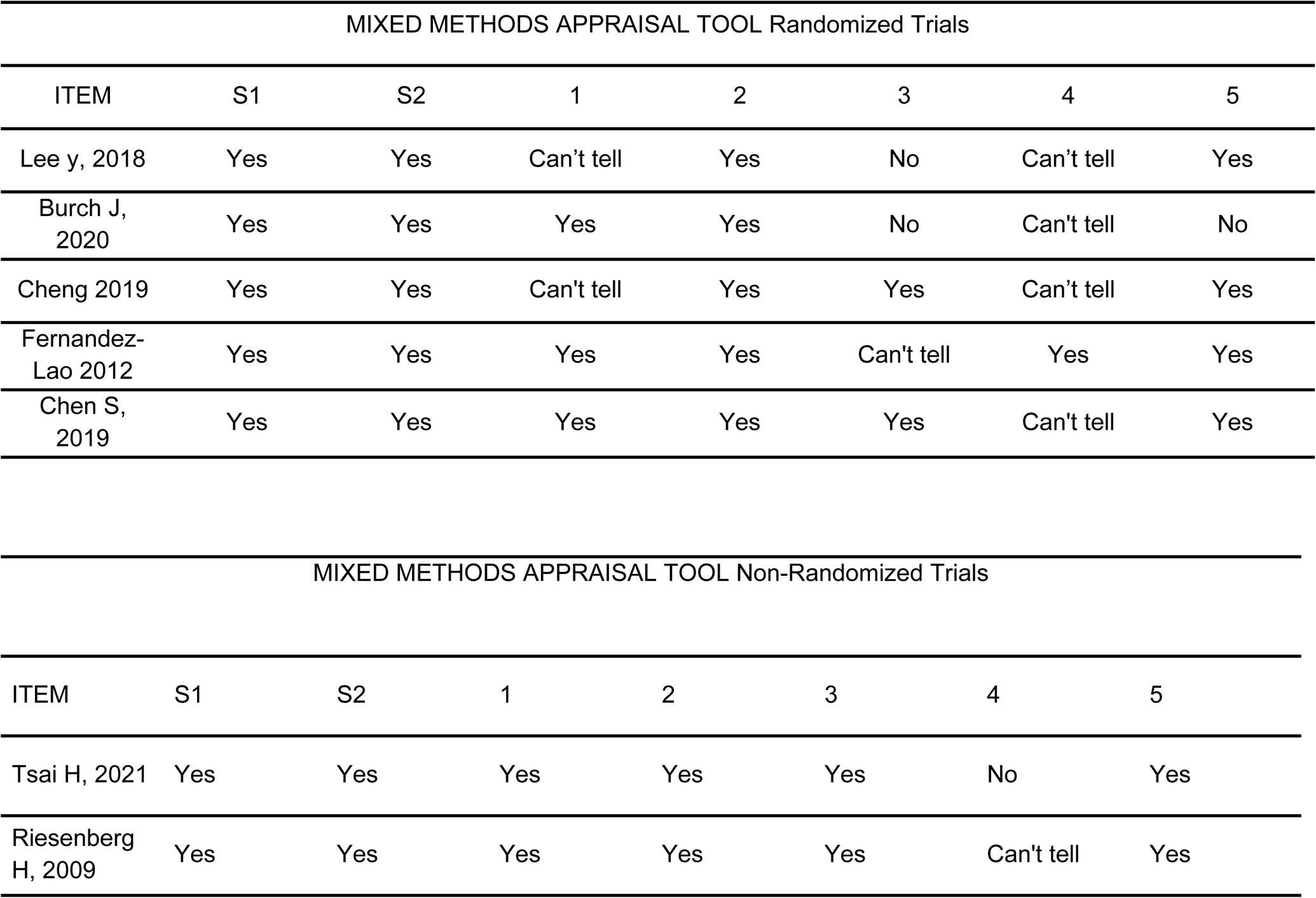
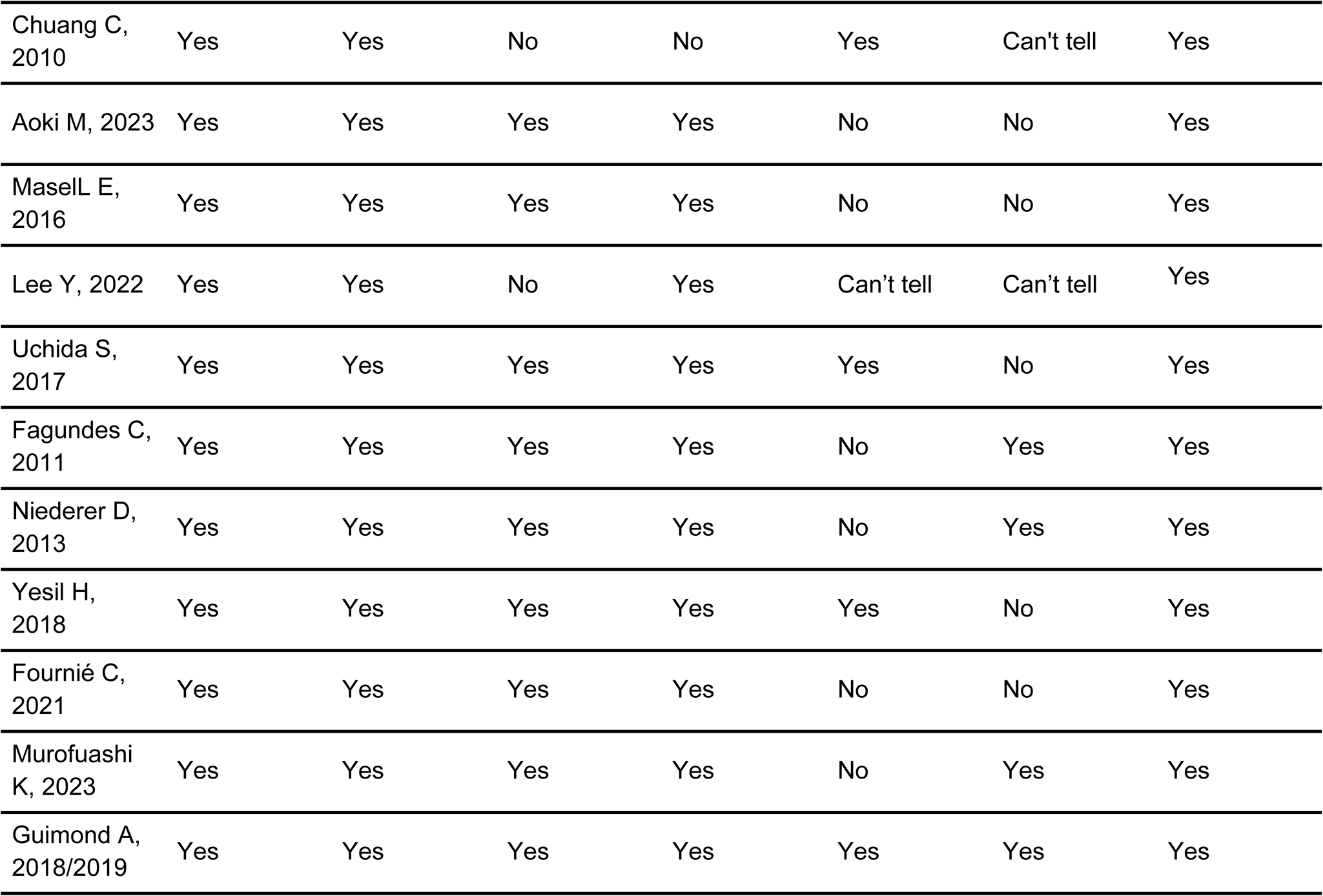
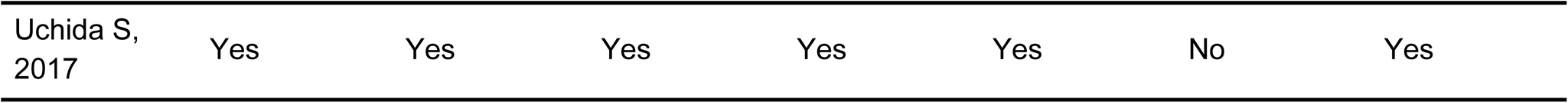
Mixed Methods Appraisal Tool Results.

